# Anti-inflammatory effects of oral prednisolone at stable state in people treated with mepolizumab: a proteomic and bulk transcriptomics analysis

**DOI:** 10.1101/2024.02.14.24302812

**Authors:** I. Howell, F. Yang, V. Brown, J. Cane, E. Marchi, A Azim, J. Busby, P.J. McDowell, S.E. Diver, C. Borg, L. G. Heaney, I. D. Pavord, C. E. Brightling, R. Chaudhuri, T.S.C. Hinks

**Affiliations:** Respiratory Medicine Unit, Experimental Medicine Division, Nuffield Department of Medicine, University of Oxford, John Radcliffe Hospital, Oxford, United Kingdom & National Institute for Health Research Oxford Biomedical Research Centre, John Radcliffe Hospital, Oxford, UK; University of Glasgow, Glasgow, UK; The National Heart and Lung Institute, Imperial college London, London, UK; Queen’s University Belfast, Belfast, UK; Clinical and Experimental Sciences, University of Southampton Faculty of Medicine, Sir Henry Wellcome Laboratories and NIHR Southampton Respiratory Biomedical Research Unit, Southampton University, Southampton, UK; Institute for Lung Health, Leicester NIHR BRC, University of Leicester, Leicester, UK

## Abstract

Mepolizumab is an anti-interleukin-5 monoclonal antibody treatment for severe eosinophilic asthma (SEA) that reduces asthma exacerbations. Residual airway inflammation on mepolizumab may lead to persistent exacerbations. Oral corticosteroids have broad anti-inflammatory effects and remain the main treatment for these residual exacerbations. Our study aimed to explore the nature and corticosteroid-responsiveness of airway inflammation after mepolizumab treatment to find potentially treatable inflammatory mechanisms.

The MAPLE trial was a multi-centre, randomized, double-blind, placebo-controlled, crossover study of 2 weeks of high-dose oral prednisolone treatment at stable state in patients treated with mepolizumab for SEA. We analysed sputum and plasma samples from the MAPLE trial using high-throughput Olink® proteomics. We also analysed plasma microRNA, sputum proteins using ELISA, and nasal mucosal bulk RNA sequencing.

In patients receiving mepolizumab, prednisolone significantly downregulated sputum proteins related to type-2 inflammation and chemotaxis including IL-4, IL-5, IL-13, CCL24, CCL26, EDN, CCL17, CCL22, OX40 receptor, FCER2, and the ST2 receptor. Prednisolone also downregulated cell adhesion molecules, prostaglandin synthases, mast cell tryptases, MMP1, MMP12, and neuroimmune mediators. Tissue repair and neutrophilic pathways were upregulated. Type-2 proteins were also downregulated in plasma, combined with IL-12, IFN-γ, and IP-10. IL-10 and amphiregulin were upregulated. In the nasal transcriptome, prednisolone suppressed genes involved in leucocyte chemotaxis, mast cell tryptase, 15-lipoxygenase and *MMP12*. By contrast, mepolizumab differentially regulated only Galectin-10 in plasma and no sputum proteins, and in nasal tissue affected genes related to cilia, keratinisation, extracellular matrix formation, and IL-4/13 signalling.

At stable state, prednisolone has broad anti-inflammatory effects on top of mepolizumab.

**Graphical abstract:** 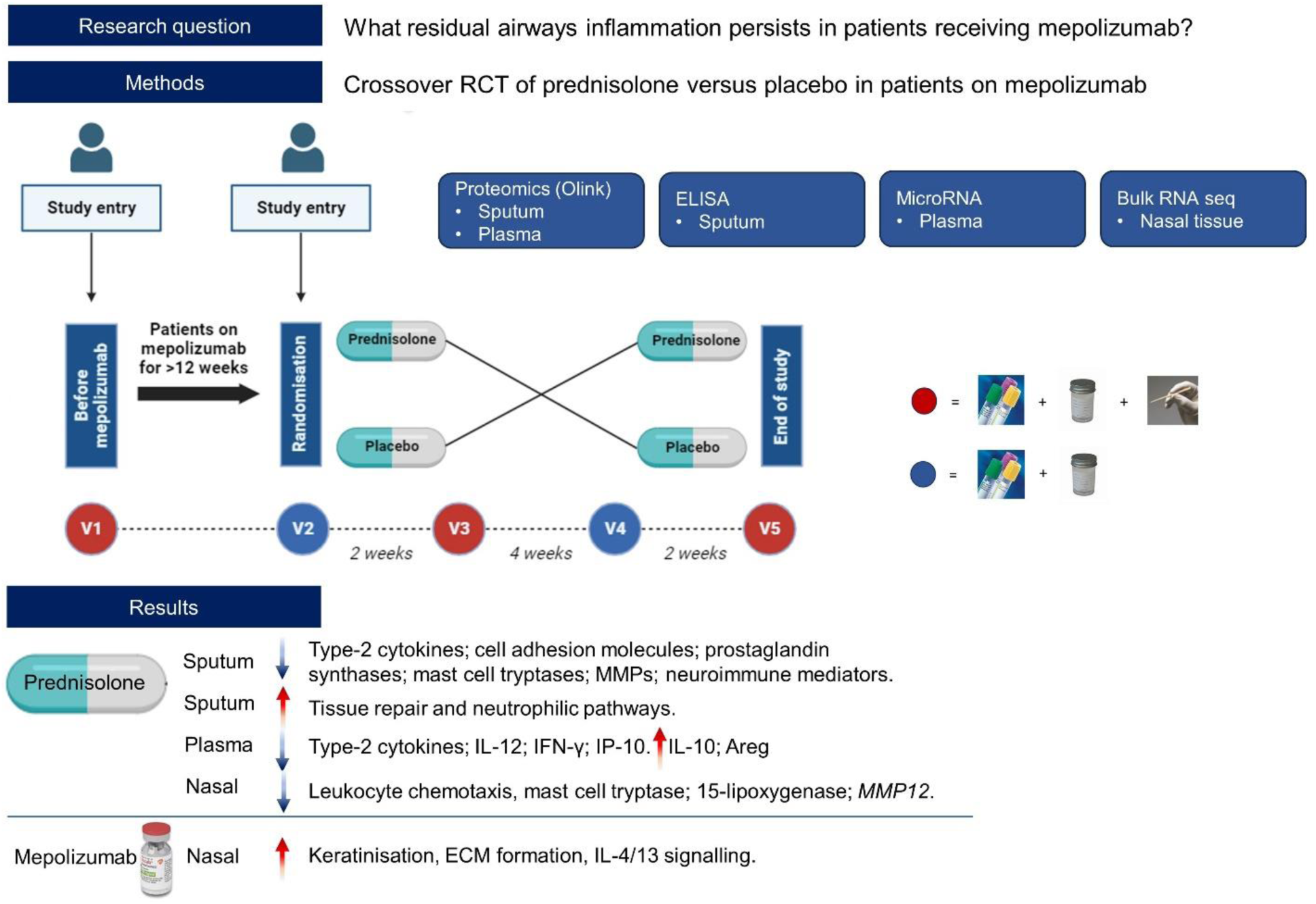

## Introduction

Oral corticosteroids (OCS) are a widely prescribed treatment for asthma exacerbations^1^. Clinical evidence for OCS at exacerbation is based on trials demonstrating a reduction in relapse and accelerated symptom resolution^2^. Biologically, OCS reduce eosinophilic inflammation, evidenced by a rapid decline in blood eosinophils through apoptosis and inhibition of interleukin-5 (IL-5)-induced production by the bone marrow^3,4^. However, OCS have broader anti-inflammatory effects that may also be clinically important. The placebo-controlled BIOAIR study identified inflammatory proteins downregulated by prednisolone in both mild/moderate and severe asthma at stable state in plasma^5^. These included chemokines and proteins involved in recruiting eosinophils to the airways^6^. But anti-inflammatory effects of OCS also extend to the airway mucosa. A placebo-controlled human bronchoscopy study in stable, moderate asthma found that prednisolone reduced airway eosinophilia, tryptase single-positive mast cells, IL-4, IL-5, and CD3+ T lymphocytes^7^.

Systemic monoclonal antibody therapy (mAb) has revolutionised the treatment of severe eosinophilic asthma. Placebo-controlled trials of mepolizumab, an anti-IL5 mAb, demonstrated approximately a 50% reduction in the annualised exacerbation rate and maintenance OCS dose^8^. This is important because repeated OCS exposure is associated with serious acute and chronic adverse effects^9^. However, some patients continue to have asthma exacerbations despite mepolizumab therapy. The MEX study showed that roughly half of these events were sputum eosinophil high, which suggests that inflammatory pathways beyond IL-5 may remain upregulated and clinically relevant in some people^10^. The nature of these pathways is not understood.

The MAPLE trial was an exploratory, multi-centre, randomized, double-blind, placebo-controlled, crossover study of prednisolone in adults treated with mepolizumab at stable state. There was no significant clinical improvement with prednisolone^11^. In this manuscript, we report the planned transcriptomic and proteomic analysis of sputum, plasma, and nasal samples from the MAPLE trial, to test the hypothesis that prednisolone exerts additional anti-inflammatory effects to mepolizumab at stable state.

## Methods

### Study design

The MAPLE study population and design were described previously^11^. Briefly, participants were recruited before or after starting mepolizumab for usual clinical care. All participants received at least 12 weeks of mepolizumab, and were OCS-free for over 4 weeks, before they were randomised (1:1) to prednisolone (0.5 mg/kg/d, maximum dose of 40 mg/d, 14 ± 2 days) followed by a 4-week washout period and then matched placebo or *vice versa* (figure 1). Prednisolone adherence was confirmed with tablet counts and serum prednisolone and cortisol levels.

**Figure 1.**
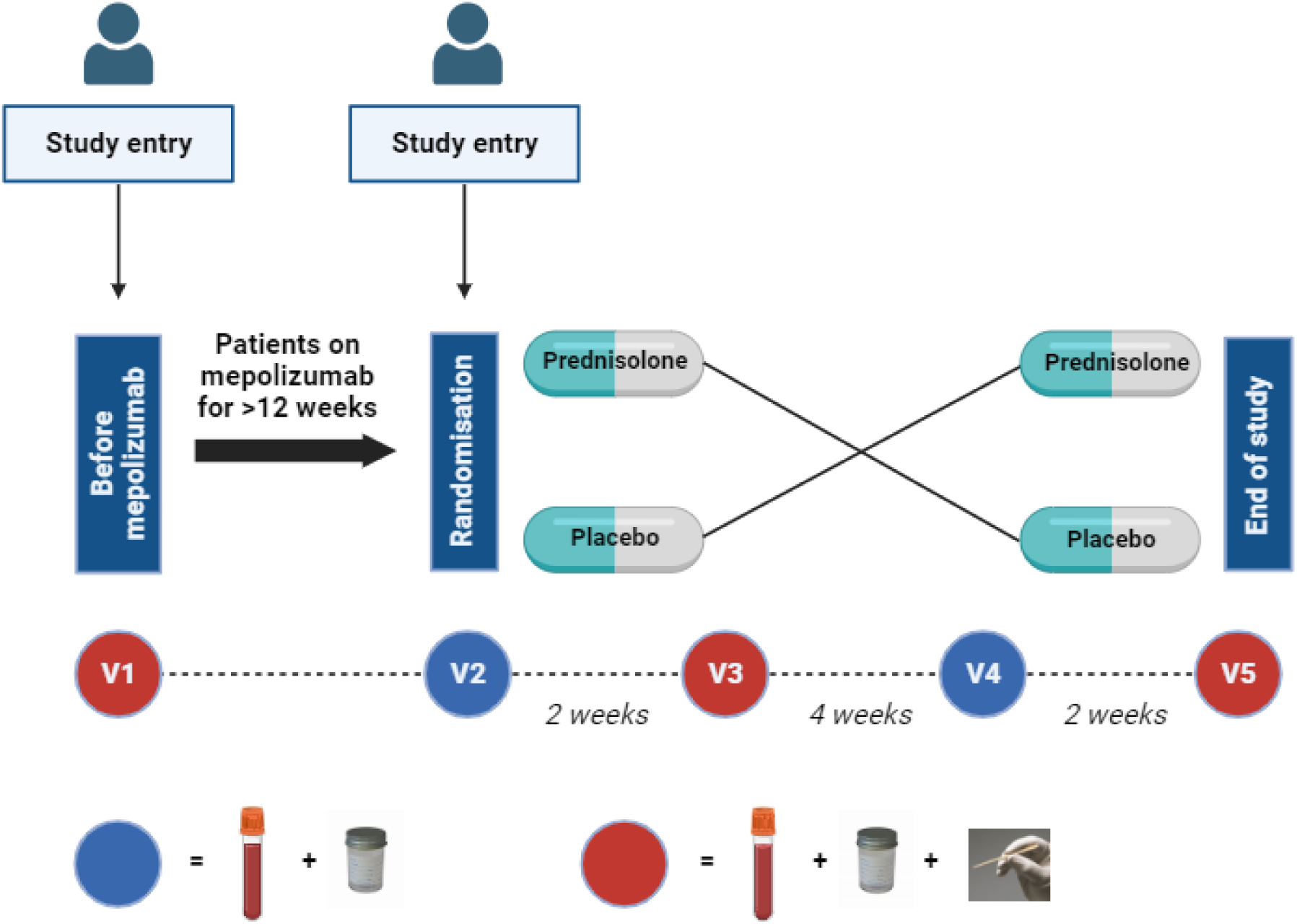
Schematic of the MAPLE study design and sampling. Participants entered the study at either visit 1 (pre-mepolizumab) or visit 2 (at least 12 weeks post-mepolizumab initiation). At visit 2 all participants were randomised to either prednisolone or placebo treatment for 2 weeks, followed by a 4-week washout, and at visit 4 crossed over to receive the opposite treatment for 2 weeks. At visits 1, 3 and 5 (red circle) participants had blood, induced sputum, and nasal scrape sampling. At visits 2 and 4 (blue circle) participants had blood and induced sputum sampling. Blood sampling consisted of EDTA and SST blood tubes.

### Sputum, blood, and nasal sampling and processing

Figure 1 describes the trial sampling.

Sputum was induced using hypertonic saline (see Appendix). Samples were transferred to the laboratory within 2 hours for initial processing. Sputum plugs were selected from saliva, weighed, and centrifuged in phosphate-buffered saline (PBS) at 790 g for 10 minutes at 4℃. Samples were aliquoted and stored at -80℃ in cryovials until further processing.

Nasal scrape samples were obtained using disposable curettes from the inferior turbinate under direct visualisation (see Appendix). Cells were stored in an RNAprotect cell reagent at -80 ℃ until processing.

### Olink® proteomic analysis of sputum and plasma samples

Sputum supernatant and plasma from all visits were analysed by Olink® Explore 1536 proximity extension assays for 1536 proteins. Olink® hybridises DNA oligonucleotide-labelled antibody pairs to proteins, extends oligonucleotides by DNA polymerase, quantifies them with next generation sequencing, and log2 transforms concentrations to a normalised protein expression (NPX) value.

Statistical analyses were performed with the Olink® Analyze package in R, according to manufacturer’s guidance. We determined the effect of prednisolone *versus* placebo on paired sputum and plasma proteins using a change from baseline analysis in a linear mixed effects model^12^. The type and sequence of treatment were fixed effects and participant ID was the random effect, with Benjamini-Hochberg (BH) correction for multiplicity of testing. An FDR <0.05 and absolute log2fold change >0.5 was considered significant. The effect of mepolizumab on sputum and serum proteins was determined using paired t-tests with BH correction. Pathway analysis was performed using Cluster Profiler, heatmaps using Pheatmap, and correlation matrices using Corrplot.

### ELISA of sputum samples

Sputum supernatants from all visits were assayed using the Ella® ELISA platform (Bio-Techne) for 15 analytes: IL-17E/IL-25, IL-1α, IL-1β, IL-2Ra, IL-33, IL-6, IL-6R, MPO, IL-3, YKL-40, IL-1RA, GM-CSF, TSLP, EDN, IL1R1. The effect of prednisolone *versus* placebo was analysed on paired samples using linear mixed effects models, and the effect of mepolizumab using paired t-tests, with BH correction (FDR<0.05).

### Bulk transcriptomics of nasal tissue samples

RNA was extracted using Qiagen RNeasy mini kits. Samples with RNA integrity number <5 and/or undetectable RNA quantity were excluded from sequencing. RNA poly A sequencing was performed by Novogene using Illumina Novaseq PE150. Gene alignment and quantification was performed by Novogene.

Differentially expressed genes were identified in DESeq2 using paired t-tests on gene count data between post-treatment placebo and prednisolone samples, and pre-mepolizumab to post-mepolizumab (post-placebo) samples. These underwent BH correction. An FDR <0.1 and absolute log2fold change >0.5 was considered significant.

### MicroRNA sequencing of plasma samples

Plasma samples underwent Illumina TruSeq Small RNA analysis, read alignment, and quantification by Q^2^ solutions®. Precursor and mature miRNA read counts were analysed for differential expression using a linear mixed effects model for the placebo *versus* prednisolone comparison, and paired t-tests comparing pre- to post-mepolizumab in DESeq2. These underwent BH correction. An FDR <0.1 and absolute log2fold change >0.5 was considered significant.

### Sample size

As this was a secondary analysis on a trial powered on clinical endpoints, a power calculation was not performed. Findings are exploratory and descriptive.

## Results

### Comparison between prednisolone and placebo

#### Olink®

16 participants (supplementary table 1 and 2) had paired sputum samples before and after prednisolone and placebo in the crossover trial for Olink® analysis. 158 proteins were significantly downregulated by prednisolone and 51 proteins were significantly upregulated (figure 2, table 1, supplementary table 3 and 4).

**Figure 2.**
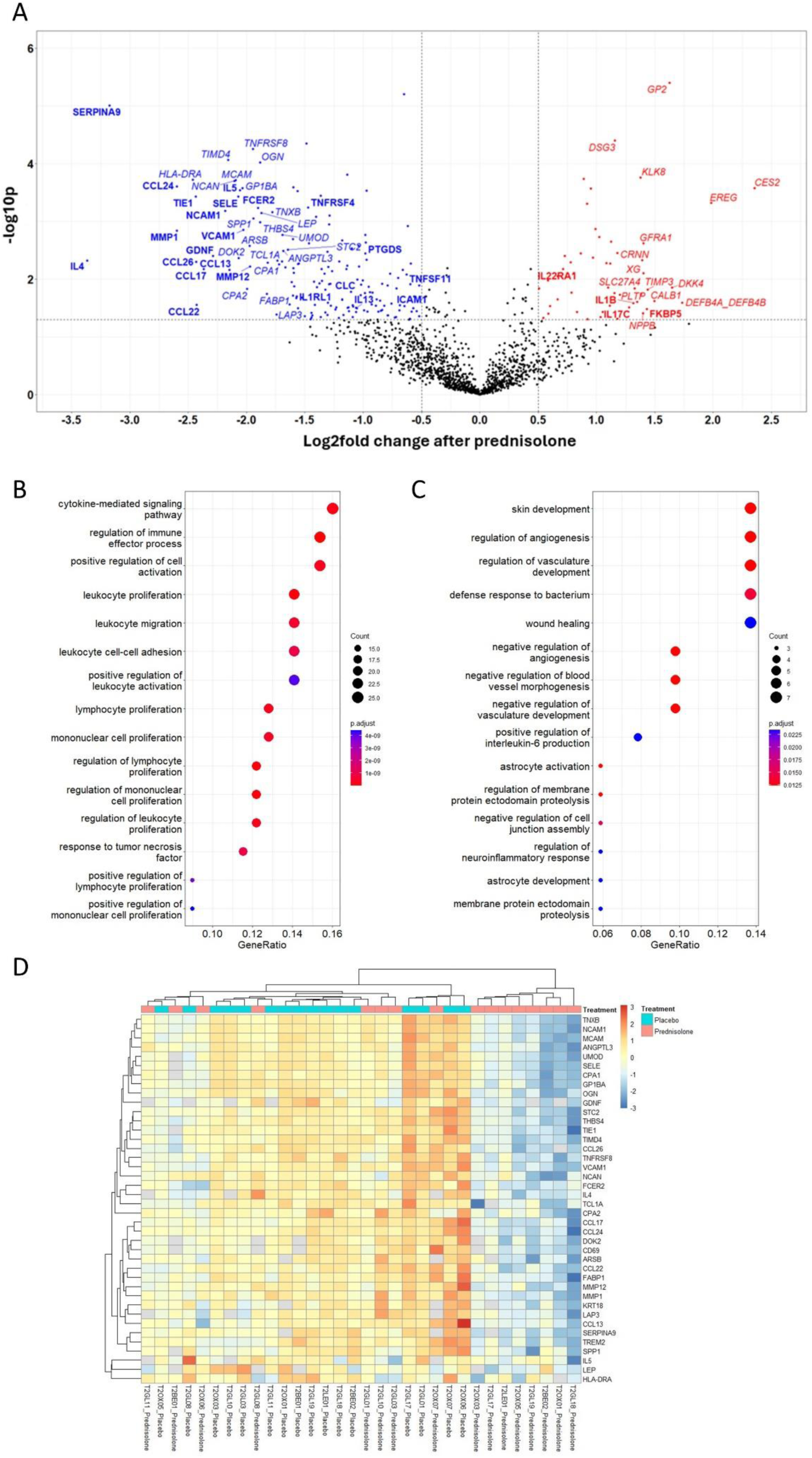
Summary of prednisolone effects on proteins measured by Olink® in sputum. A) volcano plot of differentially expressed proteins by prednisolone in sputum. An absolute log2 fold >0.5 (vertical line) and FDR <0.05 (horizonal line) were considered significant. Blue, downregulated by prednisolone; red, upregulated by prednisolone; black, non-significant. B,C) Top 15 most over-represented Gene Ontology (GO) biological processes significantly downregulated (B) or (C) upregulated by prednisolone in sputum. Colour denotes adjusted p value (FDR<0.05). D) heatmap showing individual change in NPX (n=16 participants) after prednisolone and placebo in the 40 most significantly downregulated sputum proteins from the linear mixed effects model. Participants and proteins underwent unsupervised clustering. Top row denotes treatment received, either placebo (blue) or prednisolone (red). Other rows: red denotes increased protein expression, blue reduced expression.

**Table 1.**
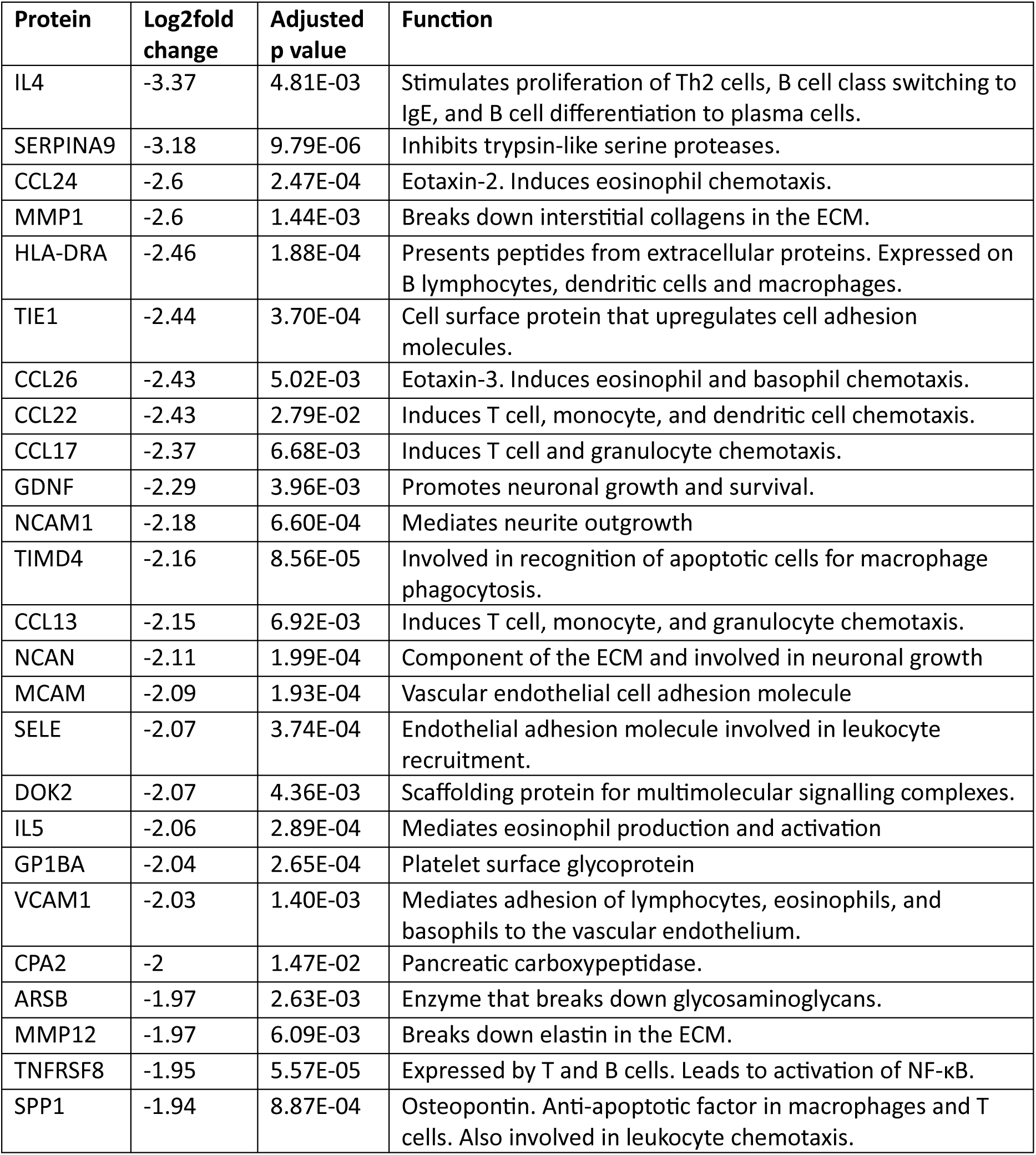
Summary of 25 most downregulated proteins by prednisolone versus placebo in sputum (Olink®)

Pathway analysis in sputum showed key downregulated biological processes including chemokine pathways crucial to subepithelial Th2/Tc2 cell, eosinophil and mast cell recruitment, and pathways related to IL-4 activity. Highly downregulated proteins related to type-2 inflammation and chemotaxis included IL-4, IL-13, IL-5, eotaxins 2 and 3 (CCL24 and CCL26), the chemokines CCL17 (TARC) and CCL22, OX40 receptor (TNFRSF4), TIMD4, the low-affinity IgE receptor FCER2, and the ST2 receptor for IL-33 (IL1RL1). Cell adhesion molecules involved in granulocyte trafficking, such as ICAM1, VCAM, TIE1, and e-selectin (SELE), were also downregulated. A reduction in the prostaglandin synthases PTGDS and HPGDS, SIGLEC6, and carboxypeptidase B1 (CPB1) that is closely related to mast cell tryptase (CPA3, not in the Olink® panel) suggests a downregulation of mast cell activity. Likewise, mast cell alpha-1/beta-1 tryptase (TPSAB1) was numerically reduced by over 50% though non-significant (adjusted p = 0.07). Proteins related to NF-kappaB (NF-κB) signalling were downregulated, including CD30 (TNFRSF8), CD69, and RANKL (TNFSF11), as were proteins related to tissue remodelling, including MMP12 and MMP1. NCAM1, NCAN, and GDNF, mediators of neuroimmune activity, were also downregulated.

Key upregulated pathways in sputum related to tissue repair and neutrophilic inflammation, such as IL-1β, IL-17C, and IL-22RA1. FKBP5, a classical hallmark of corticosteroid exposure, was upregulated.

Unsupervised clustering of the 40 proteins most downregulated in sputum after prednisolone revealed heterogeneity between individuals (n=16) in the anti-inflammatory effects of prednisolone (figure 2D). We analysed the correlation between clinical data and change in protein expression for biologically important sputum proteins after prednisolone (figure 3). This showed a reduction in sputum IL-5 and prostaglandin D synthase had an inverse correlation with FEV1, change in RANKL was correlated with change in FeNO, and IL-33 correlated with the downregulation of other type-2 cytokines and chemokines, but TSLP did not.

**Figure 3.**
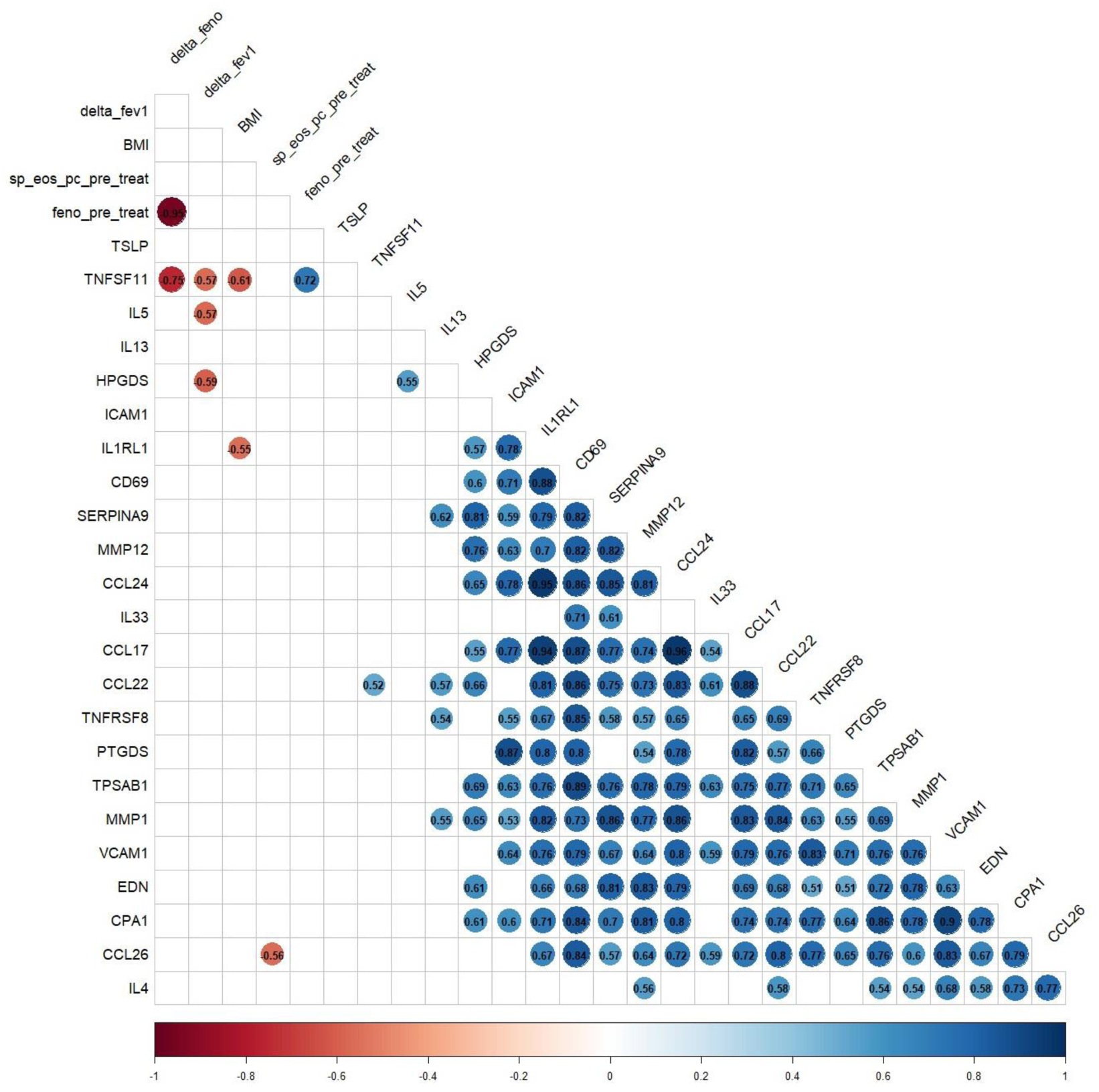
Correlation matrix of sputum proteins and clinical data. Matrix of Pearson correlation coefficients between clinical data and change in protein expression for biologically important sputum proteins after prednisolone. Only significant positive (blue) or negative (red) correlations are displayed (p<0.05). Key: feno_pre_treat, FeNO at baseline; delta_feno, change in FeNO after prednisolone; delta_fev1, change in FEV1 after prednisolone; BMI, body mass index at baseline; sp_eos_pc_pre_treatment, sputum eosinophil count at baseline

25 participants (supplementary table 1 and 2) had paired plasma samples before and after prednisolone and placebo in the crossover trial analysed by Olink®. 74 proteins were significantly downregulated by prednisolone and 53 proteins were significantly upregulated (figure 4, table 2, supplementary table 5 and 6).

**Figure 4.**
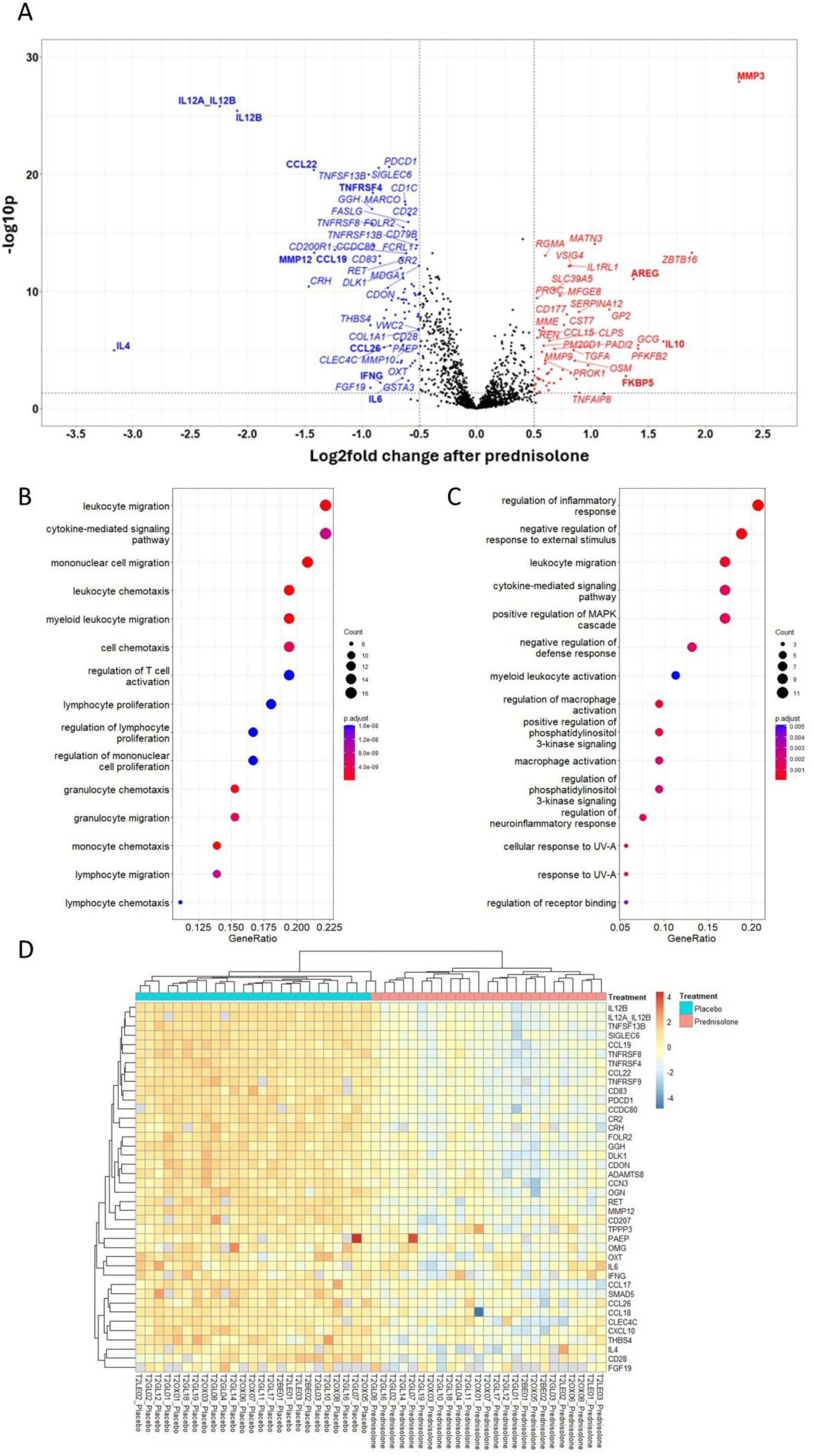
Summary of prednisolone effects on proteins measured by Olink® in plasma. A) volcano plot of differentially expressed proteins by prednisolone in plasma. An absolute log2 fold >0.5 (vertical line) and FDR <0.05 (horizonal line) were considered significant. Blue, downregulated by prednisolone; red, upregulated by prednisolone; black, non-significant. B,C) Top 15 most over-represented Gene Ontology (GO) biological processes significantly downregulated (B) or (C) upregulated by prednisolone in plasma. Colour denotes adjusted p value (FDR<0.05). D) heatmap showing individual change in NPX (n=16 participants) after prednisolone and placebo in the 40 most significantly downregulated plasma proteins from the linear mixed effects model. Participants and proteins underwent unsupervised clustering. Top row denotes treatment received, either placebo (blue) or prednisolone (red). Other rows: red denotes increased protein expression, blue reduced expression.

**Table 2.** – Summary of 25 most downregulated proteins by prednisolone versus placebo in plasma (Olink®)

| Protein | Log2fold change | Adjusted p value | Function |
| --- | --- | --- | --- |
| IL4 | -3.17 | 1.06E-05 | See table 1 |
| IL12A_IL12B | -2.24 | 1.55E-26 | Encodes a subunit of IL-12. IL-12 is produced by dendritic cells, macrophages, and neutrophils to promote Th1 differentiation. |
| IL12B | -2.09 | 3.88E-26 | Encodes a subunit of IL-12. |
| CRH | -1.46 | 3.80E-11 | Corticotrophin-releasing hormone. Stimulates ACTH production. |
| CCL22 | -1.42 | 4.18E-21 | See table 1 |
| MMP12 | -1.42 | 5.07E-14 | See table 1 |
| CCL19 | -1.24 | 2.94E-14 | Involved in T and B cell migration to secondary lymphoid organs. |
| TNFSF13B | -0.94 | 1.03E-20 | B cell activating factor. Important in B cell proliferation. |
| THBS4 | -0.93 | 7.59E-08 | Adhesive glycoprotein in cell-to-matrix interactions. |
| FGF19 | -0.93 | 1.63E-02 | Regulates bile acid synthesis. |
| GGH | -0.91 | 9.66E-18 | Key enzyme in intracellular folate homeostasis. |
| TNFRSF8 | -0.91 | 1.67E-16 | See table 1 |
| TNFRSF4 | -0.91 | 3.98E-19 | OX40 receptor. Combines with OX40 ligand to prolong T cell survival, develop memory T cells, and polarise to Th2 cells. |
| CCDC80 | -0.9 | 1.29E-14 | Promotes cell adhesion and ECM assembly. |
| SIGLEC6 | -0.86 | 2.87E-21 | Glycan-binding cell-surface protein highly expressed on mast cells. Inhibits mast cell activation after engagement with specific ligands. |
| TNFRSF9 | -0.85 | 9.89E-14 | Cell surface protein that stimulates T and B cells, and dendritic cells. |
| CD83 | -0.84 | 4.03E-13 | Stabilises MHC II. |
| IFNG | -0.84 | 6.50E-03 | Interferon gamma. Activates macrophages and MHC class II expression. |
| CLEC4C | -0.81 | 5.93E-06 | Cell surface receptor on dendritic cells involved in antigen capturing. |
| CCL18 | -0.81 | 2.03E-08 | Induces T cell, B cell, and dendritic cell chemotaxis. |
| CXCL10 | -0.79 | 3.74E-09 | Induces T cell, macrophage, and dendritic cell chemotaxis. |
| CCL17 | -0.77 | 4.81E-11 | See table 1 |
| PDCD1 | -0.77 | 2.35E-21 | Cell surface receptor on T and B cells that suppresses T cell activity. |
| CCL26 | -0.75 | 4.31E-06 | See table 1 |
| CCN3 | -0.74 | 5.80E-10 | Binds to integrin and involved ECM structure and function. |

The pathway analysis in plasma corresponded to that in sputum, showing downregulated biological processes included chemokines, type-2 inflammation pathways, and granulocyte trafficking. Relevant downregulated proteins included IL-4, eotaxin-3 (CCL26), CCL17 (TARC), CCL22, CCL19, CD83, OX40 receptor (TNFRSF4), ICAM1, VCAM. Proteins linked to remodelling such as MMP12, MMP1, and MMP10 were reduced. Proteins related to NF-κB signalling were also downregulated, including CD30 (TNFRSF8) and B-cell activating factor (TNFSF13B). There was a decrease in plasma IL-6. Additionally, plasma IL-12 activity, and associated plasma IFN-γ and IP-10 (CXCL10) levels, were significantly reduced by prednisolone.

Key upregulated processes in plasma included IL-10 and amphiregulin (AREG), important in tissue repair. MMP3 and FKBP5 were raised by prednisolone; both are previously associated with corticosteroid exposure^13,14^.

Unsupervised clustering of change in NPX after prednisolone and placebo in the 40 most downregulated plasma proteins suggested the anti-inflammatory effects of prednisolone in serum were more homogeneous compared to sputum (n=25) (figure 4D).

#### ELISA

16 participants provided sputum samples before and after prednisolone and placebo in the crossover trial that underwent ELISA. Eosinophil-derived neurotoxin (EDN) was significantly reduced after prednisolone (-1,180 ng/ml, adjusted p = 0.0001), while IL-1RA was significantly increased (16.9 ng/ml, adjusted p = 0.045). All other assays did not significantly change (supplementary figure 1, supplementary table 7).

#### Nasal tissue transcriptomics

6 participants provided good quality nasal tissue RNA post-prednisolone and post-placebo. 28 genes were significantly downregulated, and 3 genes significantly upregulated (supplementary figure 2, supplementary table 8). Key genes suppressed by prednisolone included those involved in leucocyte chemotaxis (*CXCL2, CXCL3, CX3CR1, CXCL5*), mast cell tryptase (*TPSB2*), gasdermin D (*GSDMD*), 15-lipoxygenase (*ALOX15*), and matrix metalloproteinase 12 (*MMP12*).

#### Plasma microRNA

25 participants had paired plasma tested for miRNA. There were 19 significantly differentially expressed precursor miRNAs after prednisolone. However, there were no significantly differentially expressed mature miRNAs (supplementary table 9).

### Comparison between pre-mepolizumab and post-mepolizumab

#### Olink®

Paired sputum samples from 10 participants were analysed by Olink® before and after starting mepolizumab. There were no significantly changed proteins. To understand the trend in pathways affected by mepolizumab, we performed pathway analysis using FDR <0.15 and abs log2fold >0.5. Upregulated pathways included MAPK signalling and regulation of leucocyte migration (supplementary figure 3).

Paired plasma samples from 10 participants were analysed by Olink® before and after starting mepolizumab. Galectin-10 (CLC) was the only significantly differing protein in plasma, reduced by 70%. Pathway analysis using FDR <0.15 and abs log2fold >0.2 showed mepolizumab initiation downregulated pathways related to lymphoid cell interactions and IL-3, IL-5, and GM-CSF signalling (supplementary figure 4).

#### ELISA

There were no significant differences associated with mepolizumab initiation in any of the sputum proteins tested.

#### Nasal tissue transcriptomics

5 participants provided good quality nasal tissue RNA before and after mepolizumab treatment. 272 genes were significantly downregulated and 105 significantly upregulated (supplementary table 10 and 11). In pathway analysis downregulated genes were related to ciliary structure and function. Upregulated genes were related to keratinisation, extracellular matrix formation, tight junctions, and IL-4 and IL-13 signalling pathways (figure 5).

**Figure 5.**
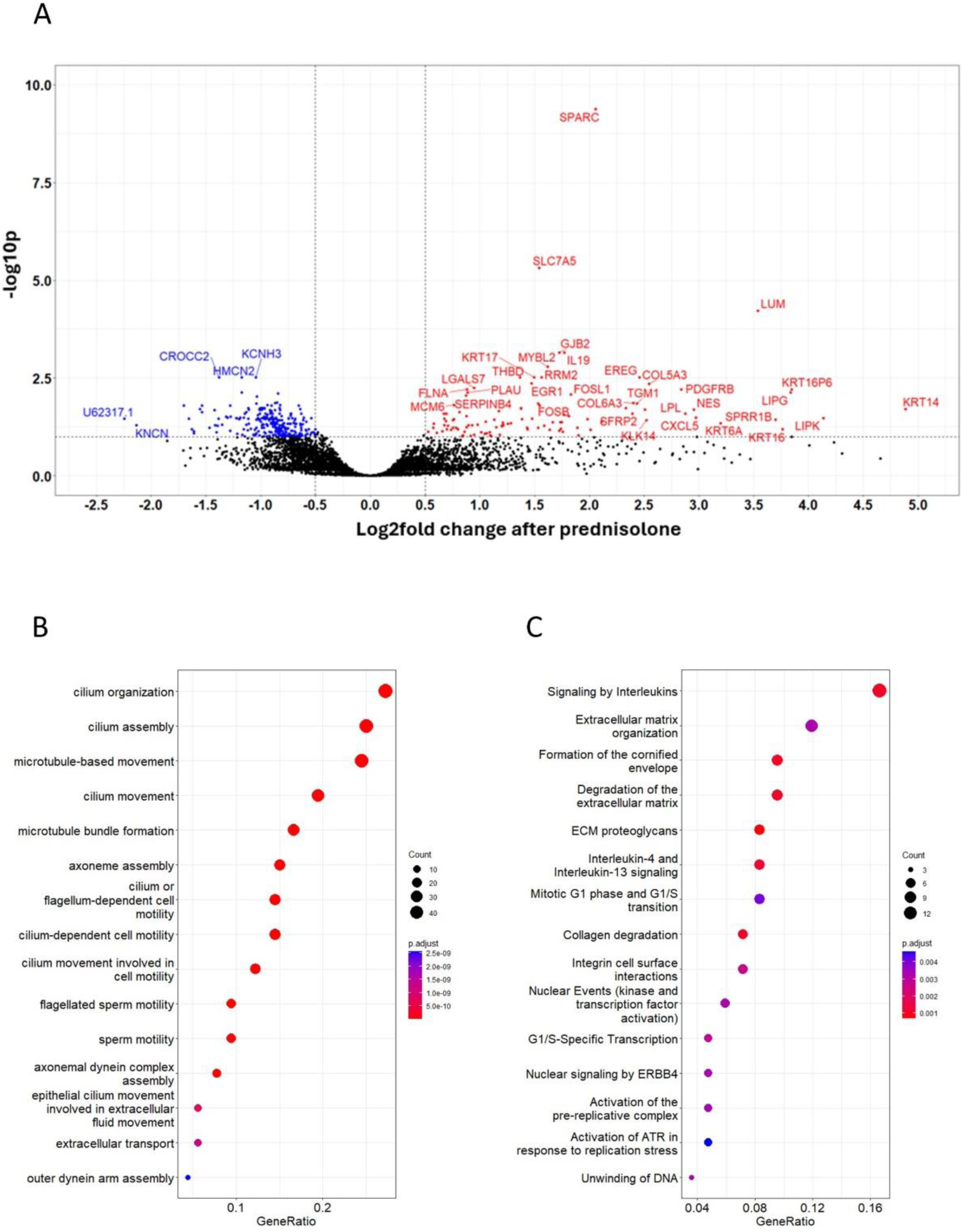
Differentially expressed nasal genes after mepolizumab. A) volcano plot of differentially expressed genes by mepolizumab in nasal tissue. An absolute log2 fold >0.5 (vertical line) and FDR <0.1 (horizonal line) were considered significant. Blue, downregulated by prednisolone; red, upregulated by prednisolone; black, non-significant. B,C) Top 15 most over-represented Gene Ontology (GO) biological processes significantly downregulated (B) or (C) upregulated by mepolizumab in nasal tissue. Colour denotes adjusted p value (FDR<0.05).

#### Plasma microRNA

There were no significant differentially expressed precursor or mature miRNAs associated with mepolizumab initiation.

## Discussion

Using a combination of high-throughput proteomics and bulk transcriptomics analysis of airway and blood samples at stable state, we found that oral prednisolone has extensive additional biological effects on top of mepolizumab.

The most striking effect of prednisolone on airway samples was the marked reduction in type-2 cytokines and chemokines (figure 6). Eotaxins, CCL17 and CCL22 are released by the airway epithelium and dendritic cells in response to IL-4 and TSLP. These chemokines have an important role in stimulating Th2/Tc2 cell and mast cell recruitment via CCR3 and CCR4 receptors. The resultant release of IL-4, IL-5, and IL-13 perpetuates Th2 cell differentiation, stimulates B cell maturation and production of IgE, and acts in concert with eotaxins, VCAM, ICAM1, E-selectin, and TIE1 to recruit eosinophils to the airway epithelium^8^. This self-perpetuating inflammation is analogous to an airway epithelial “magnet” recruiting effector granulocytes, analogous to “bombs”^15^. Our proteomic data demonstrate this process remains active and corticosteroid-responsive despite mepolizumab treatment, which selectively inhibits recruitment of eosinophils through IL-5. These findings are supported by the reduction of EDN in sputum, and CXCL2, CXCL3 and CXCL5 transcriptional suppression in nasal tissue.

**Figure 6.**
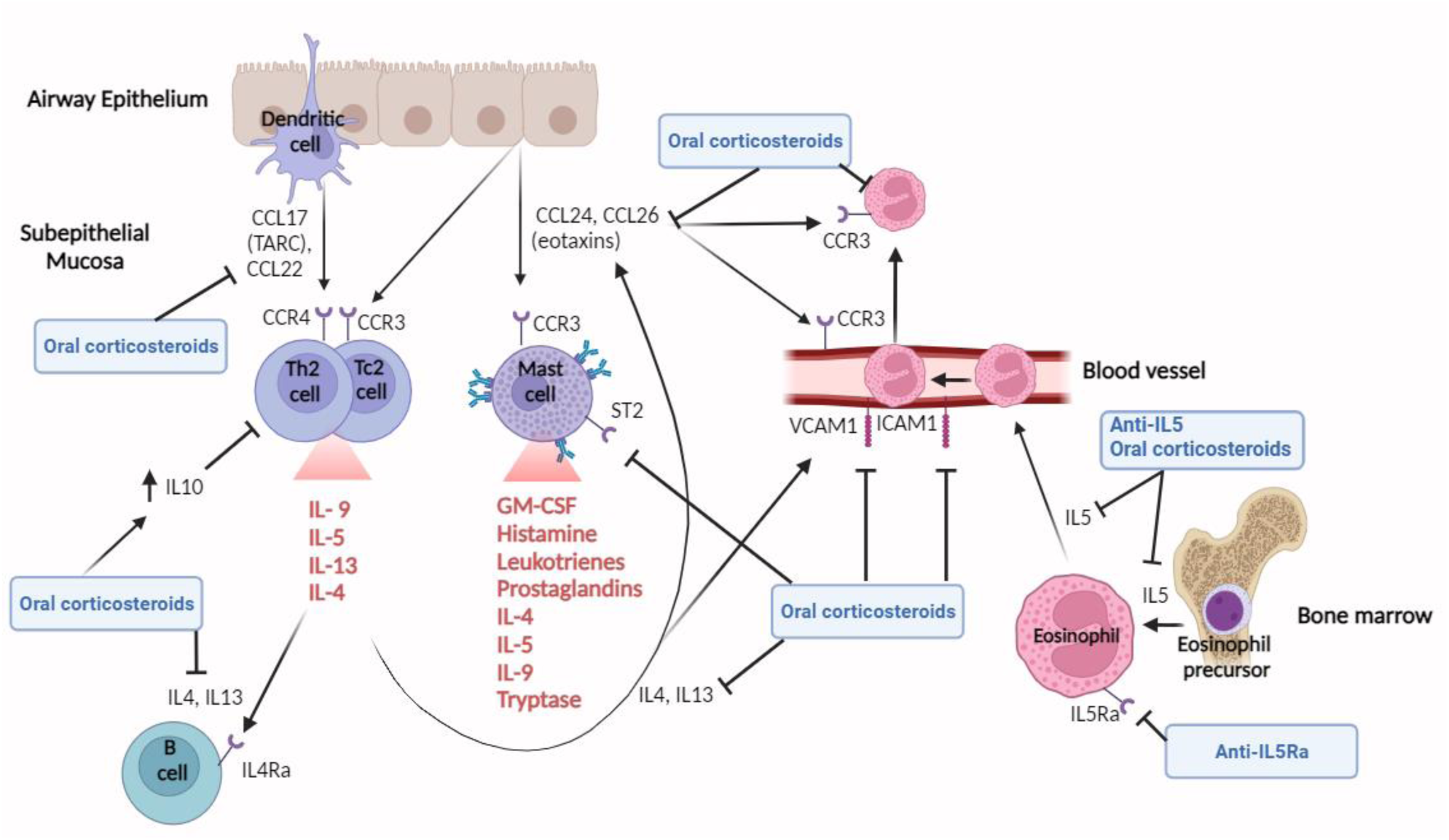
Epithelial effects of oral corticosteroids in stable asthma on mepolizumab treatment. Eotaxins (CCL24 and CCL26), CCL17 (TARC) and CCL22 are released by the airway epithelium and dendritic cells in response to IL-4 and TSLP. These chemokines have an important role in stimulating Th2/Tc2 cell and mast cell recruitment via CCR3 and CCR4 receptors. The resultant release of IL-4, IL-5, and IL-13 perpetuates Th2 cell differentiation, stimulates B cell maturation and production of IgE. IL-4 also acts in concert with eotaxins, VCAM, ICAM1, E-selectin, and TIE1 to recruit eosinophils, which are produced in the bone marrow after IL-5 stimulation, to the airway epithelium. Mepolizumab blocks IL-5, thereby reducing the production of eosinophils in the bone marrow. Oral corticosteroids have wider effects than mepolizumab, reducing IL-4, IL-13, eotaxins, CCL17, CCL22 and mast cell activity, and increasing IL-10 activity which inhibits Th2 cell production. CCL, C-C Motif Chemokine Ligand; C-C chemokine receptor, CCR; GM-CSF, Granulocyte-Macrophage Colony-Stimulating Factor; Interleukin, IL; ICAM, Intercellular Adhesion Molecule 1; Thymic Stromal Lymphopoietin, TSLP; VCAM, Vascular Cell Adhesion protein.

Biological mechanisms underpinning these effects are complex. Corticosteroids impair dendritic cell maturation, downregulate IL-4 receptor expression, inhibit IL-4/IL-13 induced STAT6 signalling, and directly inhibit e-selectin, VCAM and ICAM1 expression^16–19^. Additionally, corticosteroids upregulate the production of IL-10, as observed in our plasma proteomics, which reduces Th2 cytokine production^20^. Interestingly, the anti-inflammatory effects of prednisolone in the airway varied between individuals despite confirmed prednisolone adherence in all participants. This means that these inflammatory pathways were only active in some people at stable state, suggesting heterogeneity amongst the severe eosinophilic asthma phenotype.

Prednisolone also suppressed mast cell activity. Prostaglandin synthases (PTGDS, HPGDS) and Siglec-6 were reduced, and *15-lipooxygenase* was transcriptionally suppressed. Mast cell tryptase (CPA3) was not in the Olink® panel, but the related CPB1 which resides at the same gene locus as CPA3^21^, was significantly reduced. Additionally, the beta tryptase gene in mast cells (*TPSB2*) was transcriptionally suppressed in nasal tissue. Corticosteroids reduce mast cell signalling pathways and attenuate histamine release^22^. *Post-hoc* analysis of nasal transcriptome data in children suggested that mepolizumab may enhance mast cell and epithelial inflammation^23^. If corticosteroid-responsive mast cell activation remains present in some individuals despite mepolizumab therapy, this may be clinically relevant due to mast cell mediated airway smooth muscle contraction, and may drive some residual exacerbations which occur despite effective eosinophil depletion.

Proteomics showed prednisolone reduced the matrix metalloproteinases MMP12 and MMP1 in sputum and plasma, which correlated with type-2 epithelial cytokines. Furthermore, MMP12 transcription was significantly downregulated in nasal tissue after prednisolone. MMP12 is produced by macrophages and bronchial epithelial cells^24^. In murine models, it has been linked to airway remodelling via IL-13 dependent mechanisms^25,26^. In humans, reduced *MMP12* expression due to a functional polymorphism was associated with improved pulmonary function^27^. MMP1 expression is increased in the airways of people with asthma and is inversely associated with FEV_1_ ^28^. *In vitro*, MMP1 is induced via β3-integrin-mediated MAPK signalling leading to reduced smooth muscle contraction^29^. Additionally, MMP1 is activated by mast cell tryptase, which generates pro-proliferative extracellular matrix^30^. Our data suggest that IL-13 and mast cell-dependent MMP12 and MMP1 activation remain in some patients with asthma despite anti-IL5 treatment. The consequent potential effect on long-term lung function decline needs further investigation.

Prednisolone also significantly reduced GDNF and NCAM, mediators of neuroimmune activity, and these reductions correlated strongly with other type-2 inflammatory proteins. GDNF signals through the NCAM receptor^31^. *In vitro*, GDNF expression is increased in human airway smooth muscle cells in asthma and influences intracellular calcium responses^32^. In rat asthma models, GDNF expression was significantly higher than in controls and central sensitisation to GDNF injected in the lateral ventricle led to a significant increase in bronchial IL-4 and Th2 cells, and significant decrease in pulmonary function^33^. GDNF may contribute to smooth muscle hyperreactivity, and we found it was corticosteroid responsive.

Corticosteroids can inhibit signalling through the Toll-like receptor (TLR) cascade, including tethering and inhibiting transcription factors such as NF-κB through glucocorticoid receptors^34^. Our data suggest that components of NF-κB signalling were downregulated by prednisolone and, interestingly, this was associated with reduction in FeNO. This association is plausible because inducible nitric oxide synthase (NOS2) production is dependent on NF-κB signalling^35^. The NOS2 protein was not in the Olink® panel so we cannot comment on its response to prednisolone. The mechanisms underpinning elevated FeNO in some people despite anti-IL5 treatment is of clinical interest. There was a suggestion in our proteomic and transcriptomic pathway analysis that MAPK – which is important in TLR signalling and transactivates STAT6^36,37^ – and IL-4/IL-13 signalling pathways were upregulated following mepolizumab initiation. We also found upregulation of ciliary and extracellular matrix genes in the nasal transcriptomics that may represent epithelial repair mechanisms. This would be consistent with previous nasal transcriptome data in mepolizumab-treated children where increased expression of epithelial signalling and extracellular matrix pathways were associated with more exacerbations, likely due to increased cycles of epithelial repair post-exacerbation^38^. In adults, nasal epithelial cytokine and alarmin activation (IL-4, TSLP, IL-33) after mepolizumab treatment was associated with non-responder status^39^, suggesting a clinically relevant subgroup of severe eosinophilic asthma where inflammation is driven more by alarmins and the IL-4/13 pathway than by IL-5-mediated eosinophilia. As this epithelial signalling pathway was steroid-responsive, we speculate this inflammation could also be targeted using anti-IL4Rα, anti-TSLP, anti-IL33, or JAK inhibitors.

Our study has several strengths. Firstly, the crossover study design allowed paired comparison of biological effects of prednisolone *versus* placebo within each participant, thus avoiding inter-participant variability. Secondly, high-throughput multiomic analyses of both airway and plasma samples enabled extensive exploration of prednisolone effects in both compartments. The only previous such placebo-controlled proteomic study of prednisolone in asthma analysed only plasma^5^. Thirdly, the 4-week washout and objective measurement of plasma prednisolone levels in participants provide confidence that effects are attributable to treatment.

The main limitation of our study was modest sample size, particularly for nasal transcriptomic analysis due to low quality nasal scrape samples. Whilst the paired crossover design improved power for prednisolone *versus* placebo assessment, the lack of placebo control limited assessment of mepolizumab effects. Our findings are exploratory and should be replicated in future cohorts. Furthermore, whilst sampling occurred within a clinical trial, allowing close comparison of biological and clinical effects, our study was undertaken at stable state without long-term follow-up during exacerbations, so we did not detect significant clinical changes.

The advent of anti-IL5 monoclonal antibody therapy has dramatically altered management of severe asthma. Patients rely less on oral corticosteroids, with their extensive associated toxicity, to achieve asthma control. However, severe asthma is a heterogeneous condition. Our data demonstrate that some individuals have IL-5-independent airway epithelial inflammatory pathways which remain active and are corticosteroid-responsive. The limited effect of prednisolone on symptoms and lung function in MAPLE suggests that these residual corticosteroid responsive pathways play a minor functional role in stable patients treated with mepolizumab. However, the effect of this persistent inflammation on key asthma outcomes such as long-term lung function decline and risk of exacerbations is unknown. Prospective studies are required to determine if there are clinically relevant anti-inflammatory effects of prednisolone at exacerbation in people treated with asthma monoclonal antibodies^40^.

## Data Availability

Anonymised patient level data analysed and presented in this study are available from the corresponding author on reasonable request, providing the request meets local ethical and research governance criteria after publication. Data will be available immediately after publication for 10 years.

## Contributors

RC, LGH, IDP, CEB, FY, JB contributed to conceptualisation and design of the protocol. FY, JPM, SED, CB contributed to acquisition of study data. Lab samples were processed by IH, VB and JC. Data were analysed by IH and EM. IH and EM have accessed and verified the data. IH and TSCH drafted this submission which was approved by all authors.

## Acknowledgements

The authors are grateful to all the participants who volunteered and to the clinical and research teams at all the participating centres. For the purpose of Open Access, the author has applied a CC BY public copyright licence to any Author Accepted Manuscript version arising from this submission. The views expressed are those of the authors and not necessarily those of the NHS, the NIHR or the Department of Health and Social Care.

## Supplementary material

**Supplementary figure 1.**
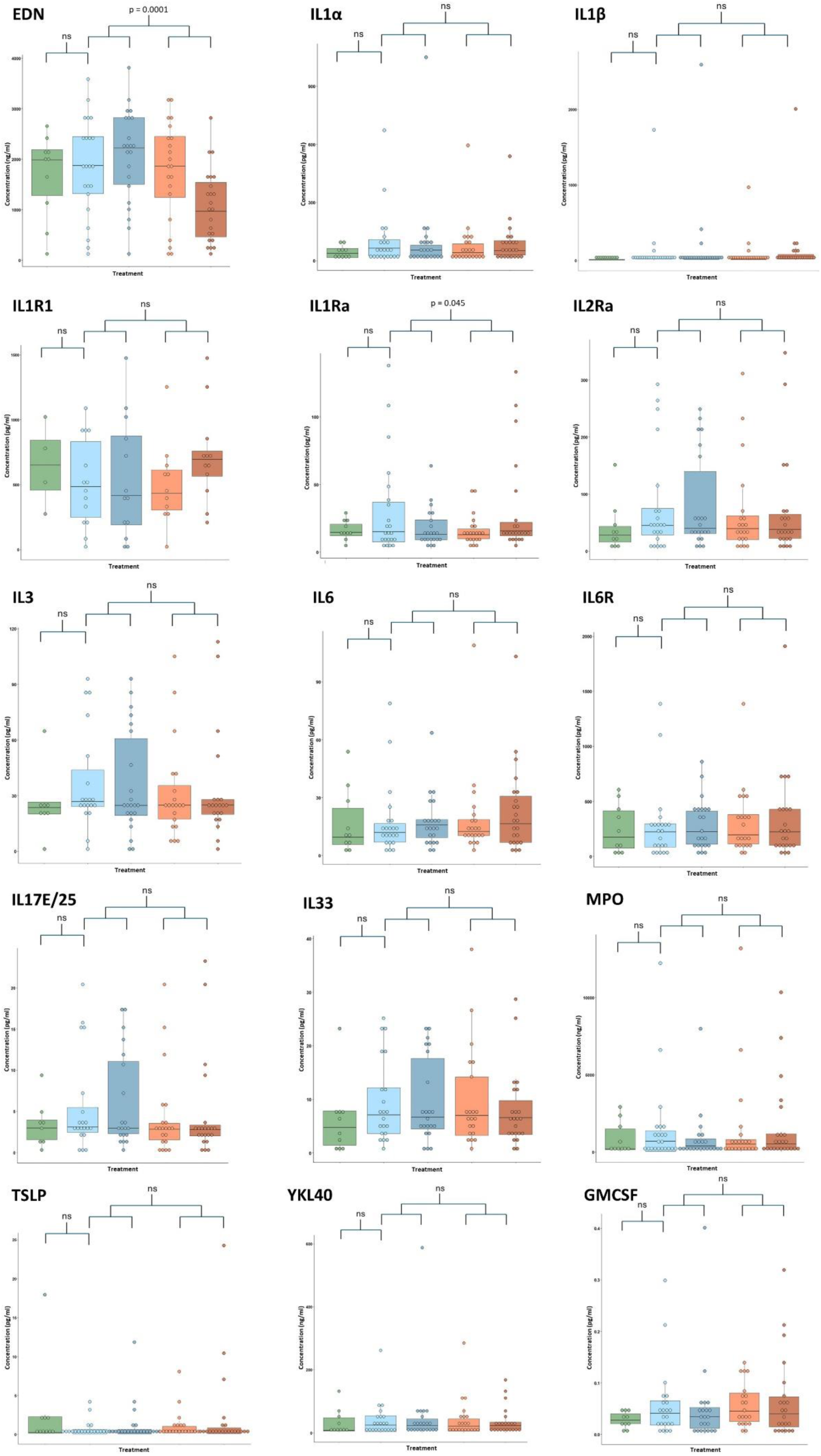
The effect of prednisolone on 15 selected proteins in sputum (ELISA) Summary of the 15 proteins tested by ELISA in sputum samples. This box and whiskers plot shows concentration versus treatment (green, before mepolizumab; light blue, before placebo; dark blue, after placebo; light red, before prednisolone; dark red, after prednisolone). The p value from before to after mepolizumab was determined using paired t-tests, and between prednisolone *versus* placebo using linear mixed effects modelling. No proteins were significantly regulated by mepolizumab. Prednisolone significantly decreased EDN, and significantly increased IL1Ra.

**Supplementary figure 2.**
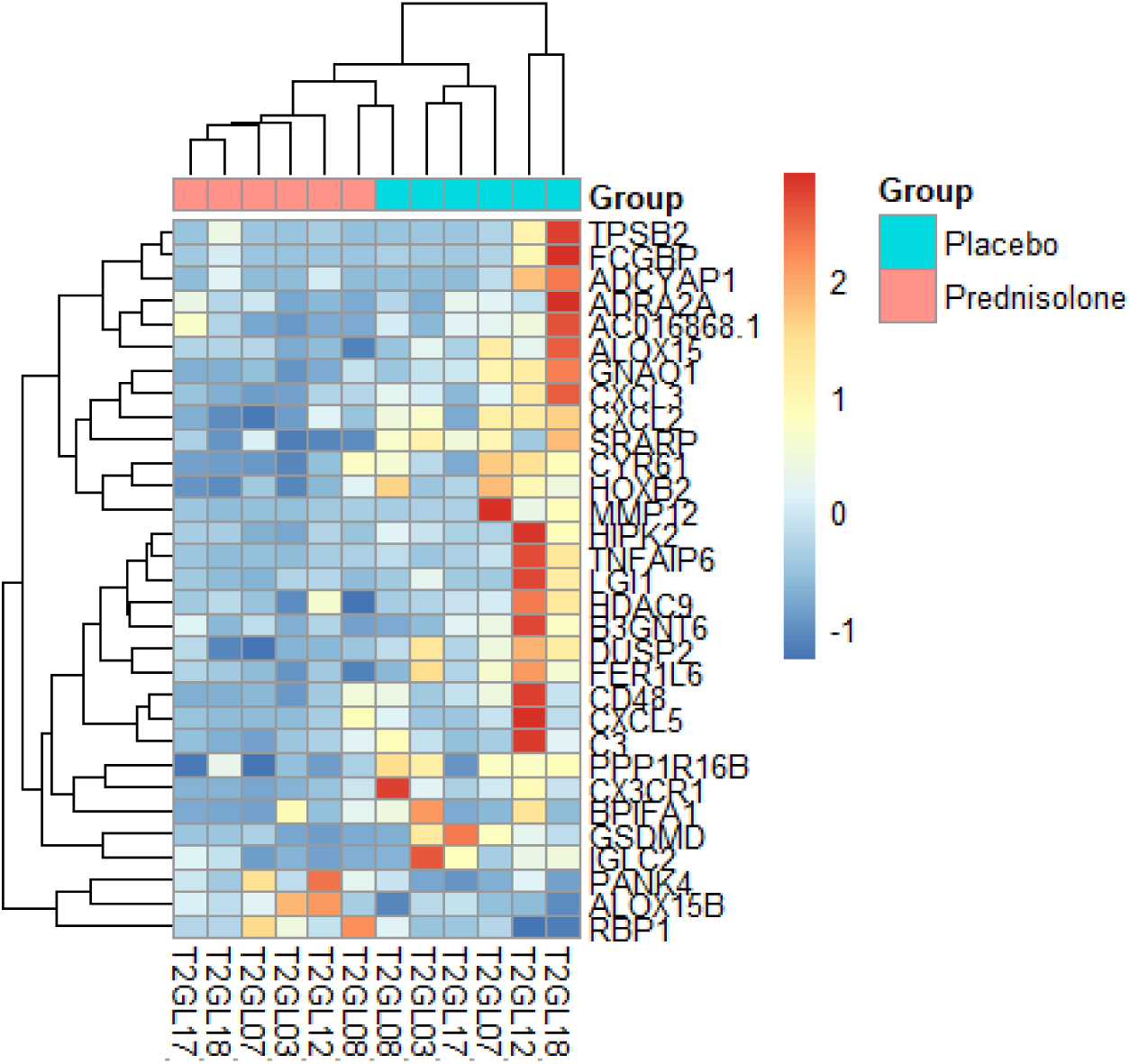
Heatmap showing effect of prednisolone and placebo on nasal transcriptome. Heatmap showing post treatment gene expression (n=6 participants) after prednisolone and placebo in the 31 significantly differentially expressed genes. Participants and genes underwent unsupervised clustering. Top row denotes treatment received, either placebo (blue) or prednisolone (red). Other rows: red denotes high gene expression, blue low expression.

**Supplementary figure 3.**
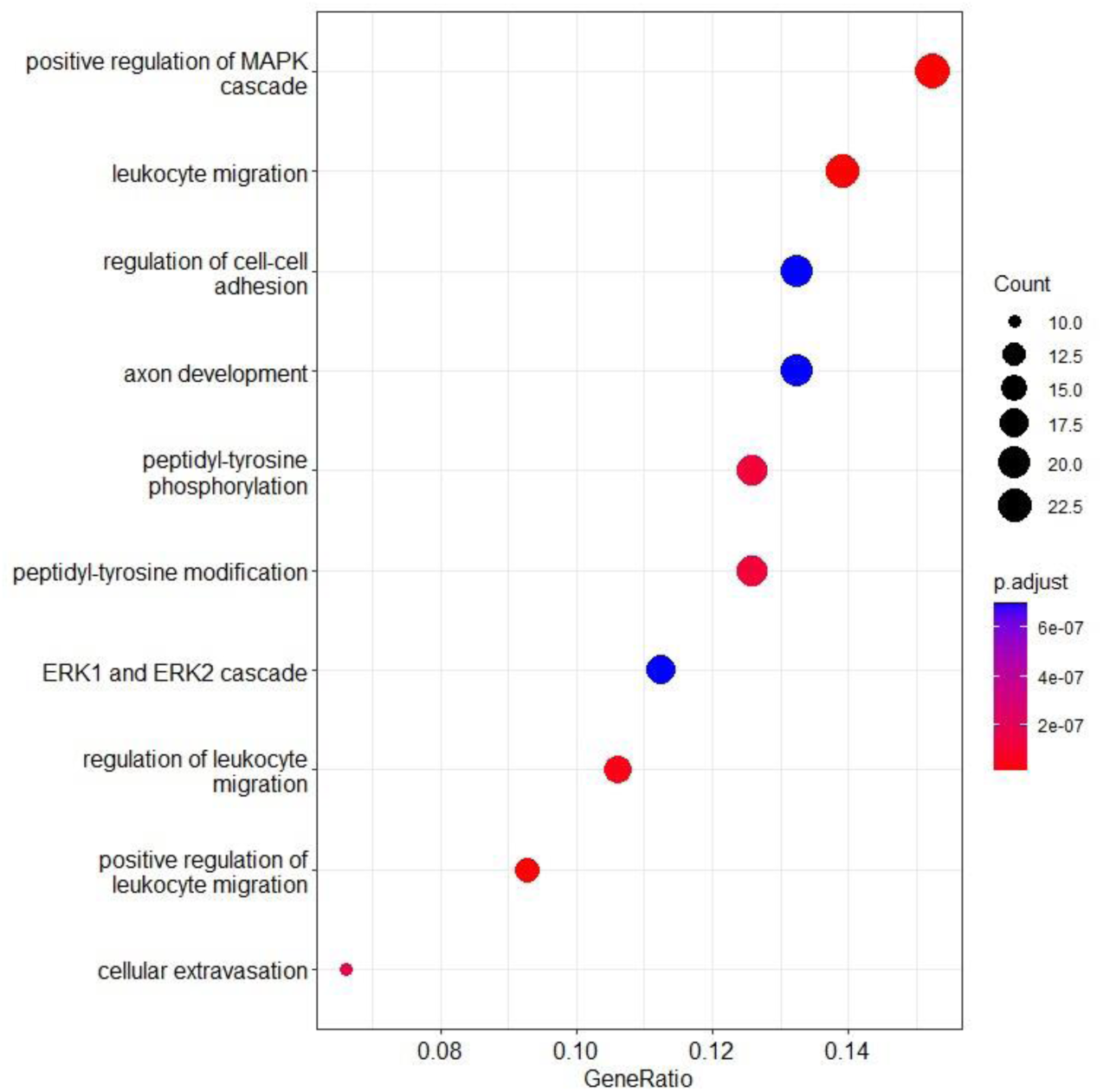
Pathways analysis of upregulated Olink® sputum proteins after mepolizumab. Top 10 most over-represented Gene Ontology (GO) biological processes for sputum proteins with a trend for upregulation by mepolizumab (FDR<0.15, abs log2fold >0.5). Colour denotes adjusted p value of pathway analysis (FDR<0.05).

**Supplementary figure 4.**
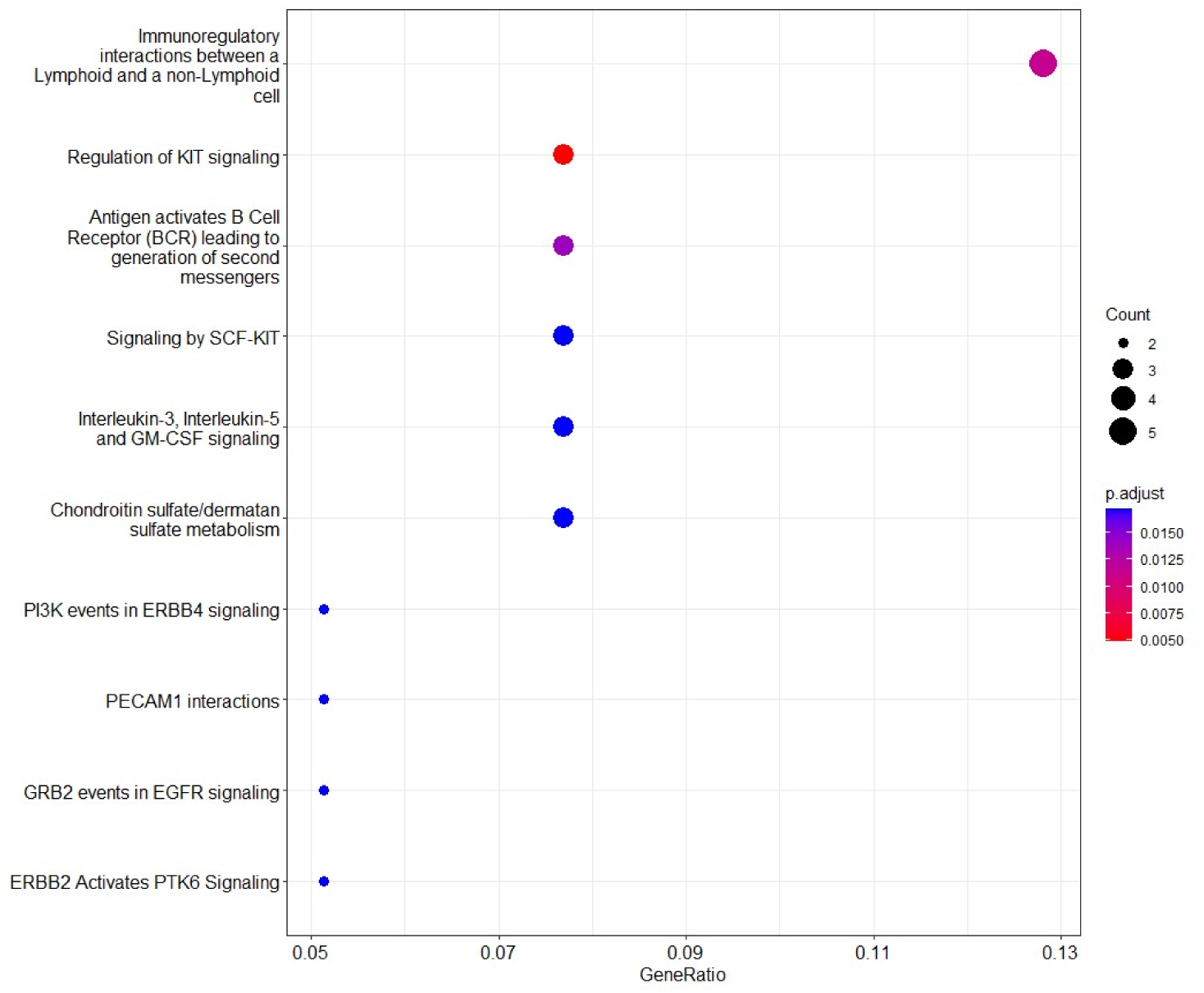
Pathways analysis of downregulated Olink® plasma proteins after mepolizumab. Top 10 most over-represented Reactome processes for plasma proteins with a trend for downregulation by mepolizumab (FDR<0.15, abs log2fold >0.2). Colour denotes adjusted p value of pathway analysis (FDR<0.05).

**Supplementary table 1.** Baseline demographics of patients who had plasma and sputum at all visits in the crossover trial compared with all patients randomised.

|  | Plasma patients | Sputum patients | All randomised patients |
| --- | --- | --- | --- |
|  | (N=25) | (N=16) | (N=27) |
| **Gender** |  |  |  |
| Female | 8 (32.0%) | 6 (37.5%) | 9 (33.3%) |
| Male | 17 (68.0%) | 10 (62.5%) | 18 (66.7%) |
| **Age Study Entry** |  |  |  |
| Mean (SD) | 56.4 (12.3) | 60.5 (8.46) | 56.9 (12.1) |
| Median [Min, Max] | 58.0 [22.0, 74.0] | 62.0 [47.0, 74.0] | 58.0 [22.0, 74.0] |
| **Ethnicity** |  |  |  |
| African | 1 (4.0%) | 1 (6.3%) | 1 (3.7%) |
| Caucasian | 24 (96.0%) | 15 (93.8%) | 26 (96.3%) |
| **Smoking Status** |  |  |  |
| Ex-smoker | 9 (36.0%) | 7 (43.8%) | 10 (37.0%) |
| Never smoker | 16 (64.0%) | 9 (56.3%) | 17 (63.0%) |
| **Pack Years** |  |  |  |
| Mean (SD) | 4.26 (9.47) | 5.53 (11.4) | 4.13 (9.14) |
| Median [Min, Max] | 0 [0, 45.0] | 0 [0, 45.0] | 0 [0, 45.0] |
| **Age Asthma Diagnosis** |  |  |  |
| Mean (SD) | 31.9 (22.2) | 39.3 (21.1) | 31.7 (22.1) |
| Median [Min, Max] | 34.0 [0, 66.0] | 43.5 [10.0, 66.0] | 34.0 [0, 66.0] |
| **BMI_v1** |  |  |  |
| Mean (SD) | 31.4 (5.38) | 32.3 (4.81) | 31.5 (5.19) |
| Median [Min, Max] | 31.9 [16.8, 40.5] | 32.2 [23.8, 40.5] | 31.9 [16.8, 40.5] |
| Missing | 1 (4.0%) | 1 (6.3%) | 1 (3.7%) |
| **Exacerbations Pre Mepo** |  |  |  |
| Mean (SD) | 0.400 (1.26) | 0.188 (0.544) | 0.370 (1.21) |
| Median [Min, Max] | 0 [0, 6.00] | 0 [0, 2.00] | 0 [0, 6.00] |
| **Exacerbations requiring ITU/HDU** |  |  |  |
| Mean (SD) | 0.160 (0.473) | 0.125 (0.342) | 0.148 (0.456) |
| Median [Min, Max] | 0 [0, 2.00] | 0 [0, 1.00] | 0 [0, 2.00] |
| **Allergic Rhinitis** |  |  |  |
| No | 21 (84.0%) | 14 (87.5%) | 22 (81.5%) |
| Yes | 4 (16.0%) | 2 (12.5%) | 5 (18.5%) |
| **Perennial Rhinitis** |  |  |  |
| No | 11 (44.0%) | 7 (43.8%) | 13 (48.1%) |
| Yes | 14 (56.0%) | 9 (56.3%) | 14 (51.9%) |
| **Nasal Polyps** |  |  |  |
| No | 17 (68.0%) | 9 (56.3%) | 18 (66.7%) |
| Yes | 8 (32.0%) | 7 (43.8%) | 9 (33.3%) |
| **Eczema** |  |  |  |
| No | 19 (76.0%) | 14 (87.5%) | 20 (74.1%) |
| Yes | 6 (24.0%) | 2 (12.5%) | 7 (25.9%) |
| **Depression/Anxiety** |  |  |  |
| No | 20 (80.0%) | 12 (75.0%) | 22 (81.5%) |
| Yes | 5 (20.0%) | 4 (25.0%) | 5 (18.5%) |
| **Bronchiectasis** |  |  |  |
| No | 23 (92.0%) | 14 (87.5%) | 24 (88.9%) |
| Yes | 2 (8.0%) | 2 (12.5%) | 3 (11.1%) |
| **Reflux** |  |  |  |
| No | 18 (72.0%) | 12 (75.0%) | 20 (74.1%) |
| Yes | 7 (28.0%) | 4 (25.0%) | 7 (25.9%) |
| **ICS Dose** |  |  |  |
| Mean (SD) | 2020 (300) | 2030 (379) | 2010 (288) |
| Median [Min, Max] | 2000 [1000, 3000] | 2000 [1000, 3000] | 2000 [1000, 3000] |
| **LABA** |  |  |  |
| Yes | 25 (100%) | 16 (100%) | 27 (100%) |
| **LTRA** |  |  |  |
| No | 9 (36.0%) | 5 (31.3%) | 9 (33.3%) |
| Yes | 16 (64.0%) | 11 (68.8%) | 18 (66.7%) |
| **LAMA** |  |  |  |
| No | 5 (20.0%) | 3 (18.8%) | 6 (22.2%) |
| Yes | 20 (80.0%) | 13 (81.3%) | 21 (77.8%) |
| **Nebuliser** |  |  |  |
| No | 22 (88.0%) | 15 (93.8%) | 24 (88.9%) |
| Yes | 3 (12.0%) | 1 (6.3%) | 3 (11.1%) |
| **Long term antibiotic** |  |  |  |
| No | 22 (88.0%) | 13 (81.3%) | 23 (85.2%) |
| Yes | 3 (12.0%) | 3 (18.8%) | 4 (14.8%) |

**Supplementary table 2.** Clinical measures at randomisation visit (V2) of patients who had plasma and sputum at all visits in the crossover trial compared with all patients randomised.

|  | Plasma patients | Sputum patients | All randomised patients |
| --- | --- | --- | --- |
|  | (N=25) | (N=16) | (N=27) |
| ACQ5 Score |  |  |  |
| N | 25 | 16 | 27 |
| Mean [SD] | 1.09 [1.05] | 1.21 [1.14] | 1.16 [1.17] |
| Median [Q1, Q3] | 0.600 [0.200, 1.80] | 0.600 [0.500, 2.05] | 0.600 [0.200, 1.90] |
| Median [Min, Max] | 0.600 [0, 3.60] | 0.600 [0, 3.60] | 0.600 [0, 4.00] |
| **mAQLQ Score** |  |  |  |
| **N** | 25 | 16 | 27 |
| **Mean [SD]** | 5.41 [1.38] | 5.41 [1.45] | 5.35 [1.48] |
| **Median [Q1, Q3]** | 5.73 [4.33, 6.53] | 5.67 [4.25, 6.63] | 5.73 [4.17, 6.57] |
| **Median [Min, Max]** | 5.73 [2.67, 7.00] | 5.67 [2.67, 7.00] | 5.73 [2.27, 7.00] |
| **SGRQ Total** |  |  |  |
| **N** | 25 | 16 | 27 |
| **Mean [SD]** | 29.8 [20.3] | 30.8 [22.0] | 30.3 [21.9] |
| **Median [Q1, Q3]** | 30.7 [14.7, 46.2] | 33.1 [15.0, 46.3] | 30.7 [13.9, 46.3] |
| **Median [Min, Max]** | 30.7 [0.850, 69.6] | 33.1 [0.850, 69.6] | 30.7 [0, 70.6] |
| **VAS Overall** |  |  |  |
| **N** | 23 | 16 | 25 |
| **Mean [SD]** | 2.47 [2.28] | 2.49 [2.34] | 2.47 [2.31] |
| **Median [Q1, Q3]** | 2.00 [0.450, 4.25] | 2.00 [0.475, 4.00] | 2.00 [0.300, 4.50] |
| **Median [Min, Max]** | 2.00 [0, 8.00] | 2.00 [0, 8.00] | 2.00 [0, 8.00] |
| **Missing** | 2 (8.0%) |  | 2 (7.4%) |
| **VAS SOB** |  |  |  |
| **N** | 23 | 16 | 25 |
| **Mean [SD]** | 2.58 [2.47] | 2.68 [2.58] | 2.65 [2.58] |
| **Median [Q1, Q3]** | 2.00 [0.400, 5.00] | 2.00 [0.500, 5.00] | 2.00 [0.200, 5.00] |
| **Median [Min, Max]** | 2.00 [0, 8.00] | 2.00 [0, 8.00] | 2.00 [0, 8.00] |
| **Missing** | 2 (8.0%) |  | 2 (7.4%) |
| **VAS Cough** |  |  |  |
| **N** | 23 | 16 | 25 |
| **Mean [SD]** | 2.29 [2.43] | 2.54 [2.68] | 2.39 [2.56] |
| **Median [Q1, Q3]** | 1.00 [0.100, 4.25] | 1.20 [0.150, 5.00] | 1.00 [0, 4.50] |
| **Median [Min, Max]** | 1.00 [0, 7.60] | 1.20 [0, 7.60] | 1.00 [0, 7.60] |
| **Missing** | 2 (8.0%) |  | 2 (7.4%) |
| **Airway oscillometry R5** |  |  |  |
| **N** | 25 | 16 | 27 |
| **Mean [SD]** | 4.01 [1.72] | 3.78 [1.58] | 4.04 [1.73] |
| **Median [Q1, Q3]** | 3.53 [2.84, 4.85] | 3.47 [2.81, 4.71] | 3.53 [2.71, 4.94] |
| **Median [Min, Max]** | 3.53 [1.48, 7.78] | 3.47 [1.48, 7.64] | 3.53 [1.48, 7.78] |
| **Airway oscillometry R5-20** |  |  |  |
| **N** | 25 | 16 | 27 |
| **Mean [SD]** | 1.00 [1.09] | 0.963 [1.06] | 1.02 [1.11] |
| **Median [Q1, Q3]** | 0.670 [0.304, 1.36] | 0.720 [0.335, 1.34] | 0.670 [0.282, 1.62] |
| **Median [Min, Max]** | 0.670 [-0.460, 3.60] | 0.720 [-0.460, 3.60] | 0.670 [-0.460, 3.60] |
| **AX** |  |  |  |
| **N** | 25 | 16 | 27 |
| **Mean [SD]** | 17.5 [18.0] | 19.2 [19.5] | 18.3 [18.8] |
| **Median [Q1, Q3]** | 7.92 [4.88, 30.8] | 9.34 [4.70, 32.8] | 7.92 [4.89, 31.3] |
| **Median [Min, Max]** | 7.92 [0.730, 57.5] | 9.34 [0.730, 57.5] | 7.92 [0.730, 57.5] |
| **Airway oscillometry X5** |  |  |  |
| **N** | 25 | 16 | 27 |
| **Mean [SD]** | -1.24 [2.48] | -1.08 [2.86] | -1.39 [2.50] |
| **Median [Q1, Q3]** | -0.870 [-2.57, -0.480] | -0.840 [-2.59, -0.475] | -1.11 [-2.61, -0.535] |
| **Median [Min, Max]** | -0.870 [-6.14, 7.71] | -0.840 [-6.14, 7.71] | -1.11 [-6.14, 7.71] |
| **FEV1 (% predicted)** |  |  |  |
| **N** | 25 | 16 | 27 |
| **Mean [SD]** | 77.9 [18.9] | 81.2 [20.7] | 77.0 [20.2] |
| **Median [Q1, Q3]** | 79.0 [67.0, 88.0] | 85.0 [70.8, 99.3] | 79.0 [66.0, 91.5] |
| **Median [Min, Max]** | 79.0 [32.0, 107] | 85.0 [32.0, 107] | 79.0 [32.0, 107] |
| **FVC (% predicted)** |  |  |  |
| **N** | 25 | 16 | 27 |
| **Mean [SD]** | 92.9 [15.6] | 96.5 [14.8] | 91.8 [16.6] |
| **Median [Q1, Q3]** | 95.0 [83.0, 104] | 101 [8.38, 108] | 95.0 [8.15, 104] |
| **Median [Min, Max]** | 95.0 [52.0, 118] | 101 [70.7, 118] | 95.0 [52.0, 118] |
| **FEV1/FVC ratio** |  |  |  |
| **N** | 25 | 16 | 27 |
| **Mean [SD]** | 0.662 [0.122] | 0.661 [0.121] | 0.659 [0.122] |
| **Median [Q1, Q3]** | 0.670 [0.650, 0.720] | 0.670 [0.650, 0.703] | 0.670 [0.650, 0.730] |
| **Median [Min, Max]** | 0.670 [0.280, 0.830] | 0.670 [0.280, 0.830] | 0.670 [0.280, 0.830] |
| **FEF (% predicted)** |  |  |  |
| **N** | 18 | 9 | 18 |
| **Mean [SD]** | 0.531 [0.245] | 0.607 [0.268] | 0.531 [0.245] |
| **Median [Q1, Q3]** | 0.510 [0.383, 0.603] | 0.520 [0.490, 0.610] | 0.510 [0.383, 0.603] |
| **Median [Min, Max]** | 0.510 [0.130, 1.17] | 0.520 [0.320, 1.17] | 0.510 [0.130, 1.17] |
| **Missing** | 7 (28.0%) | 7 (43.8%) | 9 (33.3%) |
| **PEF (L/min)** |  |  |  |
| **N** | 25 | 16 | 27 |
| **Mean [SD]** | 457 [141] | 451 [143] | 446 [145] |
| **Median [Q1, Q3]** | 425 [343, 550] | 435 [337, 572] | 424 [341, 537] |
| **Median [Min, Max]** | 425 [224, 805] | 435 [224, 705] | 424 [202, 805] |
| **FeNO (ppb)** |  |  |  |
| **N** | 25 | 16 | 27 |
| **Mean [SD]** | 47.2 [32.6] | 49.5 [34.0] | 44.9 [32.4] |
| **Median [Q1, Q3]** | 37.0 [26.0, 61.0] | 40.0 [26.3, 61.5] | 37.0 [23.0, 59.0] |
| **Median [Min, Max]** | 37.0 [3.00, 134] | 40.0 [3.00, 134] | 37.0 [3.00, 134] |
| **Blood WCC** |  |  |  |
| **N** | 25 | 16 | 27 |
| **Mean [SD]** | 6.65 [1.48] | 6.96 [1.53] | 6.51 [1.52] |
| **Median [Q1, Q3]** | 6.20 [5.70, 7.82] | 6.20 [5.85, 8.04] | 6.00 [5.55, 7.46] |
| **Median [Min, Max]** | 6.20 [4.00, 10.2] | 6.20 [5.10, 10.2] | 6.00 [4.00, 10.2] |
| **Blood Neutrophils** |  |  |  |
| **N** | 25 | 16 | 27 |
| **Mean [SD]** | 3.91 [1.10] | 4.01 [1.16] | 3.82 [1.11] |
| **Median [Q1, Q3]** | 3.70 [3.20, 4.40] | 3.75 [3.38, 4.18] | 3.70 [3.02, 4.25] |
| **Median [Min, Max]** | 3.70 [2.57, 6.51] | 3.75 [2.60, 6.51] | 3.70 [2.45, 6.51] |
| **Blood Eosinophils** |  |  |  |
| **N** | 25 | 16 | 27 |
| **Mean [SD]** | 0.0784 [0.0657] | 0.0856 [0.0756] | 0.0763 [0.0637] |
| **Median [Q1, Q3]** | 0.0600 [0.0400, 0.0900] | 0.0600 [0.0400, 0.0950] | 0.0600 [0.0400, 0.0900] |
| **Median [Min, Max]** | 0.0600 [0.0200, 0.320] | 0.0600 [0.0200, 0.320] | 0.0600 [0.0200, 0.320] |
| **Sputum Macrophages (% total)** |  |  |  |
| **N** | 20 | 15 | 22 |
| **Mean [SD]** | 24.2 [23.5] | 22.5 [21.1] | 28.8 [28.0] |
| **Median [Q1, Q3]** | 16.8 [8.16, 28.7] | 15.8 [9.45, 23.4] | 19.5 [9.08, 44.1] |
| **Median [Min, Max]** | 16.8 [1.10, 81.1] | 15.8 [1.10, 70.0] | 19.5 [1.10, 99.2] |
| **Missing** | 5 (20.0%) | 1 (6.3%) | 5 (18.5%) |
| **Sputum Neutrophils (% total)** |  |  |  |
| **N** | 20 | 15 | 22 |
| **Mean [SD]** | 64.3 [28.6] | 68.3 [25.6] | 60.6 [30.6] |
| **Median [Q1, Q3]** | 72.4 [46.9, 84.4] | 75.8 [58.2, 86.1] | 69.8 [37.1, 82.6] |
| **Median [Min, Max]** | 72.4 [12.8, 98.8] | 75.8 [17.5, 97.0] | 69.8 [0.500, 98.8] |
| **Missing** | 5 (20.0%) | 1 (6.3%) | 5 (18.5%) |
| **Sputum Eosinophils (% total)** |  |  |  |
| **N** | 20 | 15 | 22 |
| **Mean [SD]** | 6.90 [16.6] | 6.88 [18.6] | 6.29 [15.9] |
| **Median [Q1, Q3]** | 1.04 [0, 5.92] | 0.730 [0.100, 4.88] | 0.645 [0, 5.31] |
| **Median [Min, Max]** | 1.04 [0, 73.5] | 0.730 [0, 73.5] | 0.645 [0, 73.5] |
| **Missing** | 5 (20.0%) | 1 (6.3%) | 5 (18.5%) |
| **Sputum Lymphocytes (% total)** |  |  |  |
| **N** | 20 | 15 | 22 |
| **Mean [SD]** | 0.370 [0.687] | 0.476 [0.767] | 0.390 [0.669] |
| **Median [Q1, Q3]** | 0 [0, 0.275] | 0 [0, 0.515] | 0 [0, 0.325] |
| **Median [Min, Max]** | 0 [0, 2.17] | 0 [0, 2.17] | 0 [0, 2.17] |
| **Missing** | 5 (20.0%) | 1 (6.3%) | 5 (18.5%) |

**Supplementary table 3.**
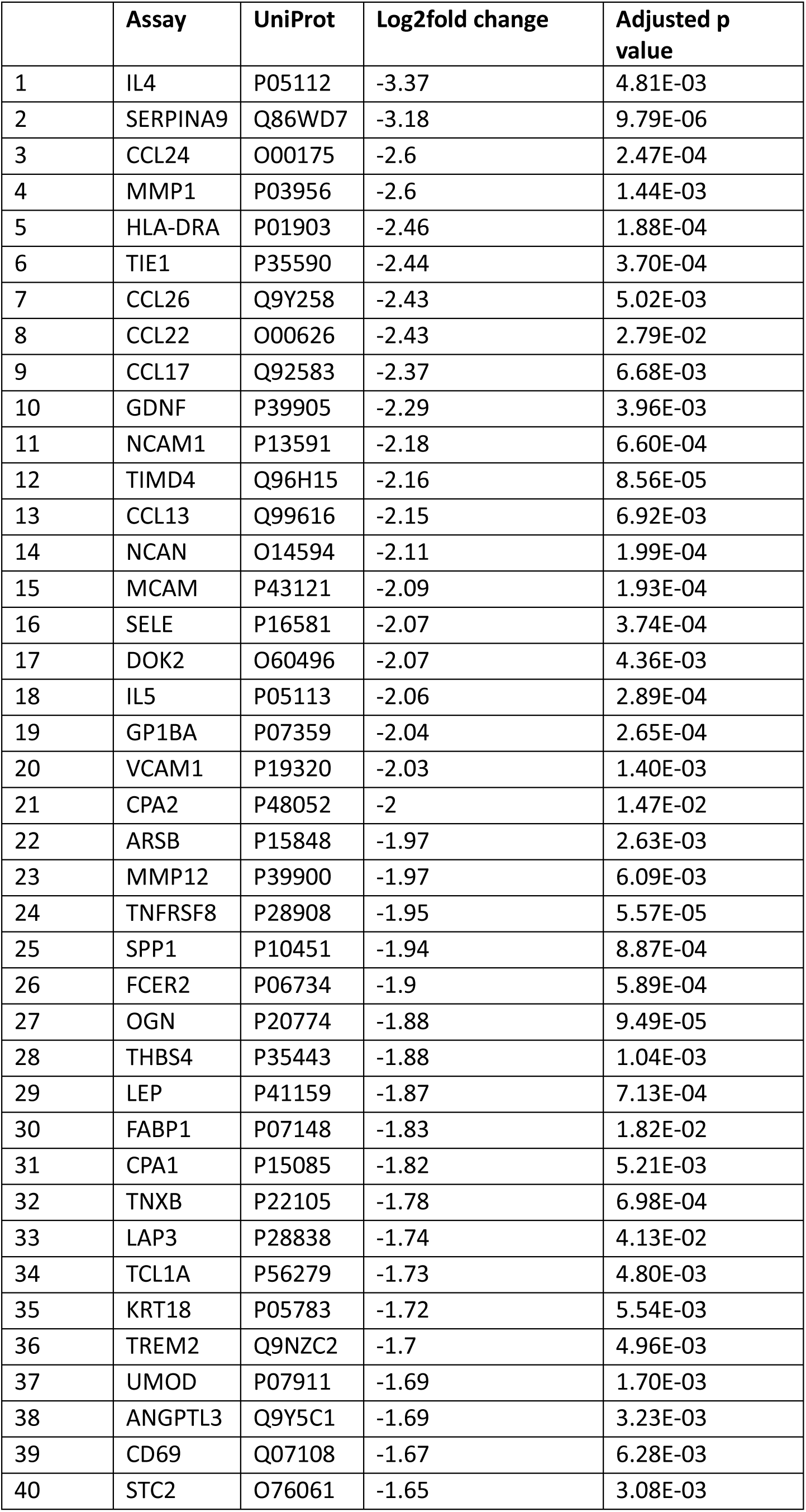
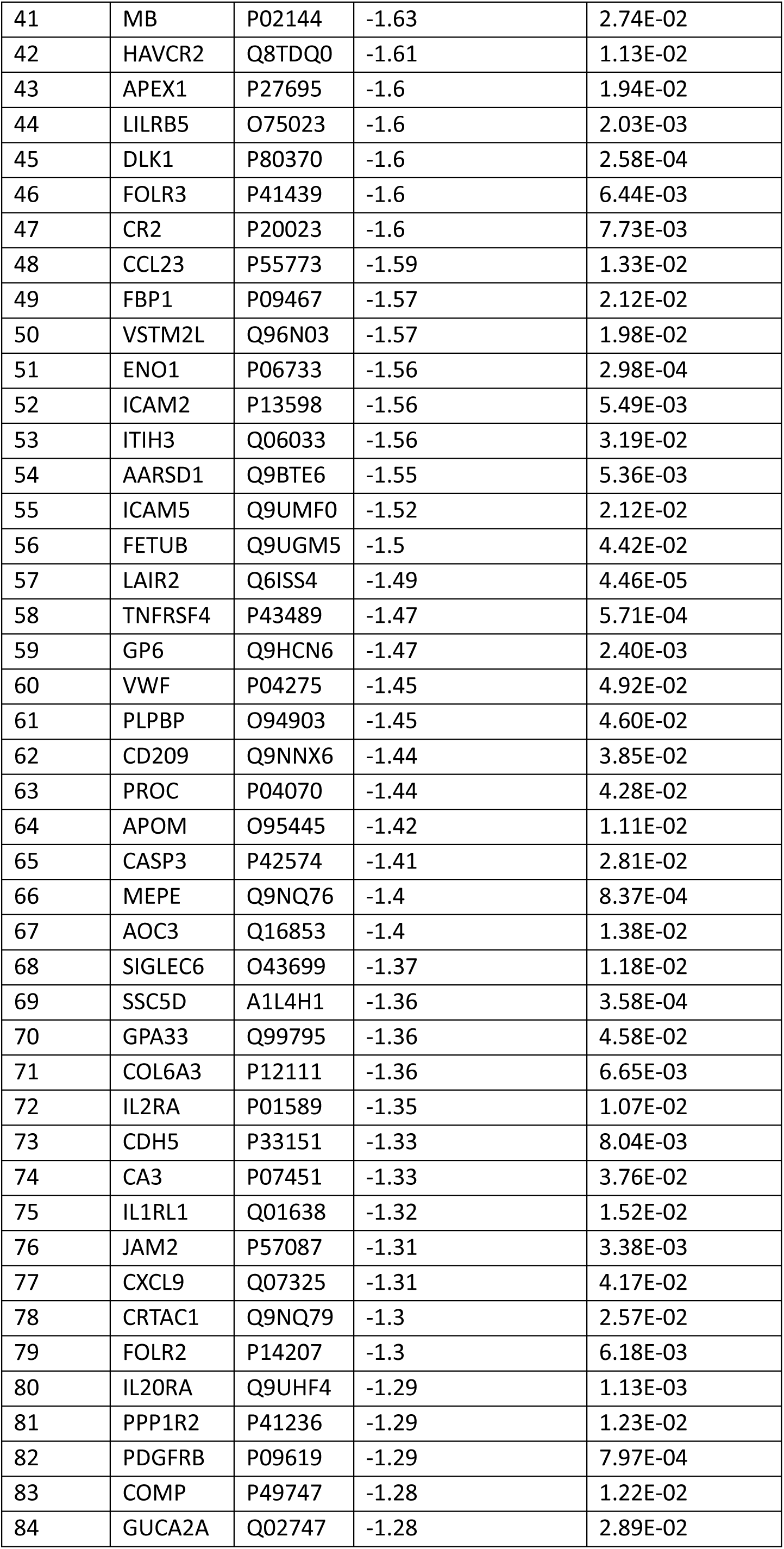
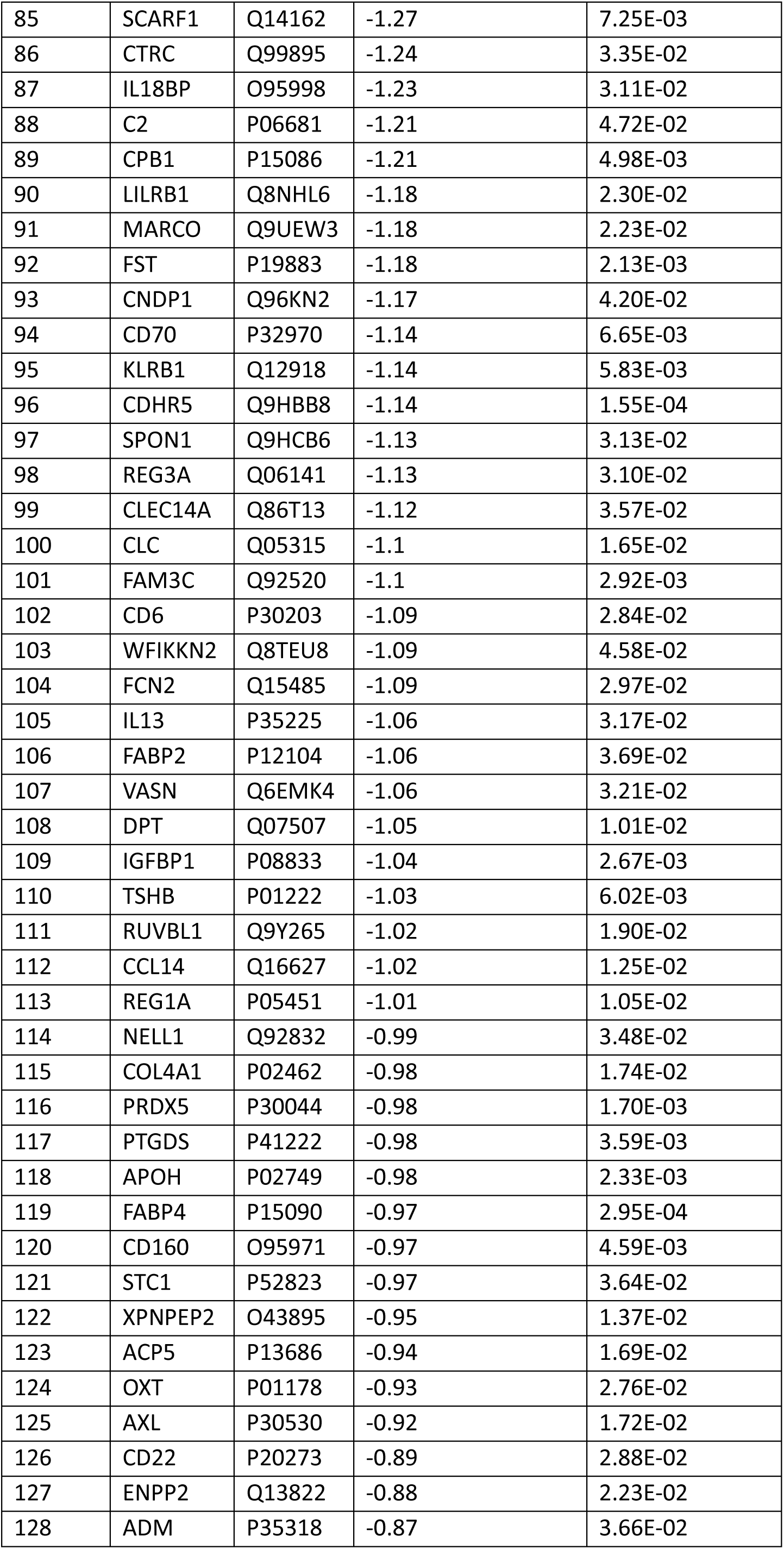
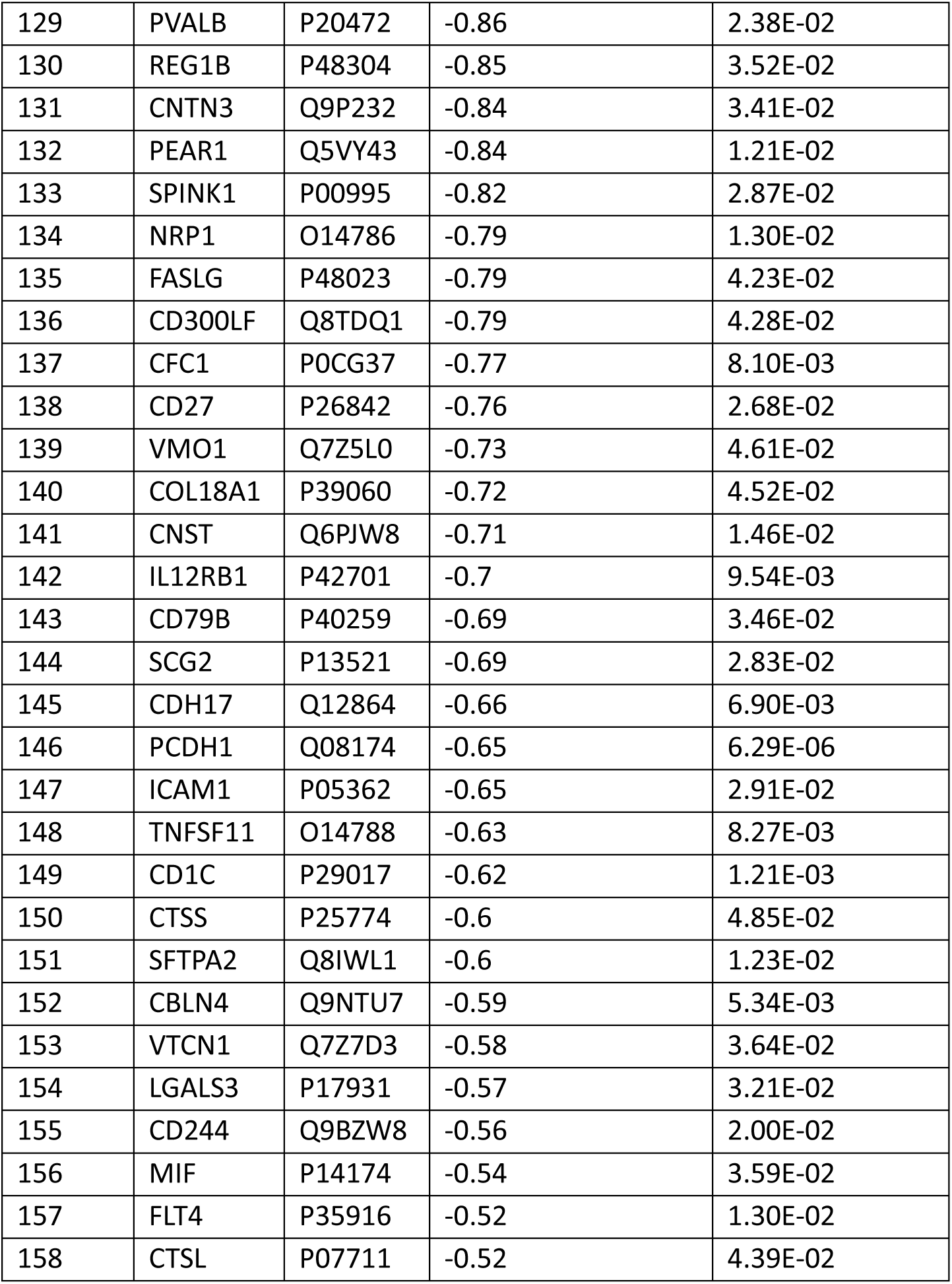
Proteins significantly downregulated by prednisolone versus placebo in sputum (Olink®)

**Supplementary table 4.** Proteins significantly upregulated by prednisolone versus placebo in sputum (Olink®)

|  | Assay | UniProt | Log2fold change | Adjusted p value |
| --- | --- | --- | --- | --- |
| 1 | CES2 | O00748 | 2.35 | 2.65E-04 |
| 2 | EREG | O14944 | 1.98 | 4.81E-04 |
| 3 | DEFB4A_DEFB4B | O15263 | 1.73 | 2.56E-02 |
| 4 | DKK4 | Q9UBT3 | 1.65 | 1.41E-02 |
| 5 | GP2 | P55259 | 1.63 | 3.95E-06 |
| 6 | CALB1 | P05937 | 1.5 | 2.40E-02 |
| 7 | TIMP3 | P35625 | 1.44 | 1.51E-02 |
| 8 | FKBP5 | Q13451 | 1.43 | 3.29E-02 |
| 9 | XG | P55808 | 1.4 | 7.96E-03 |
| 10 | GFRA1 | P56159 | 1.4 | 2.40E-03 |
| 11 | NPPB | P16860 | 1.4 | 3.89E-02 |
| 12 | CRNN | Q9UBG3 | 1.39 | 4.73E-03 |
| 13 | PLAT | P00750 | 1.38 | 1.84E-02 |
| 14 | KLK8 | O60259 | 1.38 | 1.75E-04 |
| 15 | PLTP | P55058 | 1.35 | 2.48E-02 |
| 16 | SLC27A4 | Q6P1M0 | 1.33 | 1.46E-02 |
| 17 | IL1B | P01584 | 1.32 | 2.59E-02 |
| 18 | IL17C | Q9P0M4 | 1.28 | 3.11E-02 |
| 19 | EPHA2 | P29317 | 1.2 | 1.88E-02 |
| 20 | NDRG1 | Q92597 | 1.18 | 3.52E-03 |
| 21 | ARID4B | Q4LE39 | 1.18 | 4.88E-02 |
| 22 | DSG3 | P32926 | 1.16 | 4.00E-05 |
| 23 | PRSS27 | Q9BQR3 | 1.15 | 1.75E-02 |
| 24 | ANXA5 | P08758 | 1.13 | 2.24E-03 |
| 25 | CNTN5 | O94779 | 1.12 | 5.36E-03 |
| 26 | SNCG | O76070 | 1.11 | 2.89E-02 |
| 27 | ULBP2 | Q9BZM5 | 1.1 | 1.40E-02 |
| 28 | WFDC12 | Q8WWY7 | 1.08 | 5.30E-03 |
| 29 | CEACAM21 | Q3KPI0 | 1.05 | 3.74E-02 |
| 30 | FCAR | P24071 | 1.03 | 4.54E-02 |
| 31 | DSC2 | Q02487 | 1.02 | 1.88E-03 |
| 32 | SPINK5 | Q9NQ38 | 0.99 | 1.37E-03 |
| 33 | ADGRG2 | Q8IZP9 | 0.97 | 4.27E-03 |
| 34 | F2R | P25116 | 0.95 | 2.71E-04 |
| 35 | CA12 | O43570 | 0.92 | 4.93E-02 |
| 36 | KLK6 | Q92876 | 0.92 | 4.97E-04 |
| 37 | IVD | P26440 | 0.89 | 1.82E-04 |
| 38 | CDSN | Q15517 | 0.87 | 3.62E-03 |
| 39 | LDLR | P01130 | 0.83 | 3.69E-02 |
| 40 | DNER | Q8NFT8 | 0.81 | 7.91E-03 |
| 41 | PSMA1 | P25786 | 0.78 | 1.73E-02 |
| 42 | ESM1 | Q9NQ30 | 0.78 | 5.11E-03 |
| 43 | CHMP1A | Q9HD42 | 0.73 | 3.95E-03 |
| 44 | HS3ST3B1 | Q9Y662 | 0.72 | 6.73E-03 |
| 45 | IL22RA1 | Q8N6P7 | 0.67 | 8.90E-03 |
| 46 | DCN | P07585 | 0.67 | 2.24E-02 |
| 47 | PODXL2 | Q9NZ53 | 0.6 | 2.95E-02 |
| 48 | ACVRL1 | P37023 | 0.59 | 3.96E-02 |
| 49 | CTSV | O60911 | 0.58 | 1.05E-02 |
| 50 | KCNIP4 | Q6PIL6 | 0.54 | 4.69E-02 |
| 51 | PSG1 | P11464 | 0.51 | 1.44E-02 |

**Supplementary table 5.** Proteins significantly downregulated by prednisolone versus placebo in plasma (Olink®)

|  | Assay | UniProt | Log2fold change | Adjusted p value |
| --- | --- | --- | --- | --- |
| 1 | IL4 | P05112 | -3.17 | 1.06E-05 |
| 2 | IL12A_IL12B | P29459_P29460 | -2.24 | 1.55E-26 |
| 3 | IL12B | P29460 | -2.09 | 3.88E-26 |
| 4 | CRH | P06850 | -1.46 | 3.80E-11 |
| 5 | CCL22 | O00626 | -1.42 | 4.18E-21 |
| 6 | MMP12 | P39900 | -1.42 | 5.07E-14 |
| 7 | CCL19 | Q99731 | -1.24 | 2.94E-14 |
| 8 | TNFSF13B | Q9Y275 | -0.94 | 1.03E-20 |
| 9 | THBS4 | P35443 | -0.93 | 7.59E-08 |
| 10 | FGF19 | O95750 | -0.93 | 1.63E-02 |
| 11 | GGH | Q92820 | -0.91 | 9.66E-18 |
| 12 | TNFRSF8 | P28908 | -0.91 | 1.67E-16 |
| 13 | TNFRSF4 | P43489 | -0.91 | 3.98E-19 |
| 14 | CCDC80 | Q76M96 | -0.9 | 1.29E-14 |
| 15 | SIGLEC6 | O43699 | -0.86 | 2.87E-21 |
| 16 | TNFRSF9 | Q07011 | -0.85 | 9.89E-14 |
| 17 | CD83 | Q01151 | -0.84 | 4.03E-13 |
| 18 | IFNG | P01579 | -0.84 | 6.50E-03 |
| 19 | CLEC4C | Q8WTT0 | -0.81 | 5.93E-06 |
| 20 | CCL18 | P55774 | -0.81 | 2.03E-08 |
| 21 | CXCL10 | P02778 | -0.79 | 3.74E-09 |
| 22 | CCL17 | Q92583 | -0.77 | 4.81E-11 |
| 23 | PDCD1 | Q15116 | -0.77 | 2.35E-21 |
| 24 | CCL26 | Q9Y258 | -0.75 | 4.31E-06 |
| 25 | CCN3 | P48745 | -0.74 | 5.80E-10 |
| 26 | CD207 | Q9UJ71 | -0.71 | 9.97E-09 |
| 27 | OXT | P01178 | -0.69 | 1.11E-04 |
| 28 | ADAMTS8 | Q9UP79 | -0.68 | 4.01E-10 |
| 29 | CR2 | P20023 | -0.66 | 1.47E-13 |
| 30 | TPPP3 | Q9BW30 | -0.66 | 1.30E-04 |
| 31 | DLK1 | P80370 | -0.66 | 1.06E-12 |
| 32 | CD28 | P10747 | -0.66 | 1.73E-06 |
| 33 | OMG | P23515 | -0.65 | 8.41E-05 |
| 34 | OGN | P20774 | -0.65 | 4.84E-10 |
| 35 | RET | P07949 | -0.65 | 2.20E-13 |
| 36 | IL6 | P05231 | -0.65 | 2.51E-03 |
| 37 | CDON | Q4KMG0 | -0.64 | 7.29E-12 |
| 38 | PAEP | P09466 | -0.64 | 3.66E-06 |
| 39 | FOLR2 | P14207 | -0.64 | 3.48E-16 |
| 40 | SMAD5 | Q99717 | -0.64 | 5.05E-10 |
| 41 | CCL21 | O00585 | -0.64 | 9.54E-10 |
| 42 | CD70 | P32970 | -0.64 | 1.23E-10 |
| 43 | CD300E | Q496F6 | -0.63 | 6.27E-09 |
| 44 | ITIH3 | Q06033 | -0.63 | 2.09E-08 |
| 45 | CD1C | P29017 | -0.62 | 2.26E-18 |
| 46 | LAMP3 | Q9UQV4 | -0.62 | 6.56E-11 |
| 47 | MARCO | Q9UEW3 | -0.62 | 3.73E-18 |
| 48 | DRAXIN | Q8NBI3 | -0.62 | 5.19E-10 |
| 49 | CD200R1 | Q8TD46 | -0.62 | 5.60E-14 |
| 50 | MMP10 | P09238 | -0.61 | 8.08E-06 |
| 51 | FASLG | P48023 | -0.6 | 1.20E-16 |
| 52 | GSTA3 | Q16772 | -0.6 | 4.09E-03 |
| 53 | CXCL17 | Q6UXB2 | -0.59 | 1.16E-09 |
| 54 | CDCP1 | Q9H5V8 | -0.59 | 5.35E-09 |
| 55 | CD22 | P20273 | -0.59 | 3.33E-17 |
| 56 | GZMB | P10144 | -0.58 | 2.44E-04 |
| 57 | MSR1 | P21757 | -0.57 | 2.37E-11 |
| 58 | SERPINA9 | Q86WD7 | -0.56 | 1.39E-04 |
| 59 | CNTN2 | Q02246 | -0.55 | 3.55E-11 |
| 60 | LAG3 | P18627 | -0.54 | 1.43E-10 |
| 61 | IFNL1 | Q8IU54 | -0.54 | 8.64E-05 |
| 62 | KLK14 | Q9P0G3 | -0.54 | 1.79E-09 |
| 63 | LTA | P01374 | -0.54 | 2.19E-10 |
| 64 | FCRL1 | Q96LA6 | -0.53 | 2.29E-14 |
| 65 | CD79B | P40259 | -0.53 | 3.69E-15 |
| 66 | TNFRSF13B | O14836 | -0.52 | 1.20E-14 |
| 67 | ADAM23 | O75077 | -0.52 | 1.43E-08 |
| 68 | CXCL11 | O14625 | -0.51 | 2.10E-05 |
| 69 | DPT | Q07507 | -0.51 | 1.86E-10 |
| 70 | LRRC25 | Q8N386 | -0.51 | 8.36E-09 |
| 71 | COL1A1 | P02452 | -0.51 | 1.98E-07 |
| 72 | COL6A3 | P12111 | -0.51 | 1.39E-08 |
| 73 | VWC2 | Q2TAL6 | -0.5 | 1.71E-07 |
| 74 | MDGA1 | Q8NFP4 | -0.5 | 6.67E-13 |

**Supplementary table 6.** Proteins significantly upregulated by prednisolone versus placebo in plasma (Olink®)

|  | Assay | UniProt | Log2fold change | Adjusted p value |
| --- | --- | --- | --- | --- |
| 1 | MMP3 | P08254 | 2.29 | 1.21E-28 |
| 2 | ZBTB16 | Q05516 | 1.88 | 5.02E-14 |
| 3 | IL10 | P22301 | 1.63 | 1.98E-06 |
| 4 | GCG | P01275 | 1.41 | 3.93E-06 |
| 5 | PFKFB2 | O60825 | 1.41 | 7.54E-06 |
| 6 | AREG | P15514 | 1.37 | 8.78E-12 |
| 7 | FKBP5 | Q13451 | 1.3 | 1.72E-03 |
| 8 | GP2 | P55259 | 1.16 | 3.41E-09 |
| 9 | MATN3 | O15232 | 1.03 | 9.59E-15 |
| 10 | TMPRSS15 | P98073 | 0.98 | 2.01E-04 |
| 11 | TNFAIP8 | O95379 | 0.9 | 4.60E-02 |
| 12 | SERPINA12 | Q8IW75 | 0.89 | 5.98E-09 |
| 13 | AGR3 | Q8TD06 | 0.87 | 3.16E-03 |
| 14 | OSM | P13725 | 0.86 | 8.28E-05 |
| 15 | TGFA | P01135 | 0.85 | 1.10E-05 |
| 16 | STAT5B | P51692 | 0.82 | 9.60E-04 |
| 17 | VSIG4 | Q9Y279 | 0.82 | 6.29E-13 |
| 18 | IL1RL1 | Q01638 | 0.81 | 7.26E-13 |
| 19 | CD177 | Q8N6Q3 | 0.79 | 9.72E-09 |
| 20 | CST7 | O76096 | 0.76 | 7.01E-08 |
| 21 | RBP2 | P50120 | 0.75 | 5.11E-04 |
| 22 | NAMPT | P43490 | 0.74 | 3.06E-03 |
| 23 | MFGE8 | Q08431 | 0.73 | 2.38E-10 |
| 24 | PADI2 | Q9Y2J8 | 0.68 | 8.22E-06 |
| 25 | IL17C | Q9P0M4 | 0.68 | 6.68E-03 |
| 26 | SLC39A5 | Q6ZMH5 | 0.67 | 7.27E-11 |
| 27 | CEACAM8 | P31997 | 0.65 | 4.45E-04 |
| 28 | CLPS | P04118 | 0.64 | 1.69E-06 |
| 29 | PIK3AP1 | Q6ZUJ8 | 0.63 | 6.57E-03 |
| 30 | IPCEF1 | Q8WWN9 | 0.63 | 2.71E-02 |
| 31 | MMP8 | P22894 | 0.63 | 8.89E-04 |
| 32 | SCP2 | P22307 | 0.61 | 1.17E-03 |
| 33 | PADI4 | Q9UM07 | 0.61 | 1.09E-02 |
| 34 | PROK1 | P58294 | 0.6 | 8.48E-05 |
| 35 | RGMA | Q96B86 | 0.6 | 8.78E-14 |
| 36 | CES1 | P23141 | 0.6 | 1.43E-04 |
| 37 | PM20D1 | Q6GTS8 | 0.59 | 4.24E-06 |
| 38 | HCLS1 | P14317 | 0.58 | 4.08E-03 |
| 39 | MME | P08473 | 0.58 | 1.34E-07 |
| 40 | MMP9 | P14780 | 0.57 | 1.44E-05 |
| 41 | ARG1 | P05089 | 0.55 | 3.25E-03 |
| 42 | CCL20 | P78556 | 0.55 | 4.47E-02 |
| 43 | CCL15 | Q16663 | 0.55 | 2.16E-07 |
| 44 | RASSF2 | P50749 | 0.55 | 3.90E-02 |
| 45 | LEP | P41159 | 0.54 | 4.29E-04 |
| 46 | CEP43 | O95684 | 0.54 | 3.13E-03 |
| 47 | REN | P00797 | 0.53 | 9.54E-07 |
| 48 | MVK | Q03426 | 0.53 | 2.45E-03 |
| 49 | PROC | P04070 | 0.53 | 3.80E-10 |
| 50 | YTHDF3 | Q7Z739 | 0.52 | 8.40E-03 |
| 51 | FEN1 | P39748 | 0.51 | 2.17E-02 |
| 52 | BAIAP2 | Q9UQB8 | 0.51 | 1.57E-03 |
| 53 | DNAJB1 | P25685 | 0.5 | 1.15E-02 |

**Supplementary table 7.**
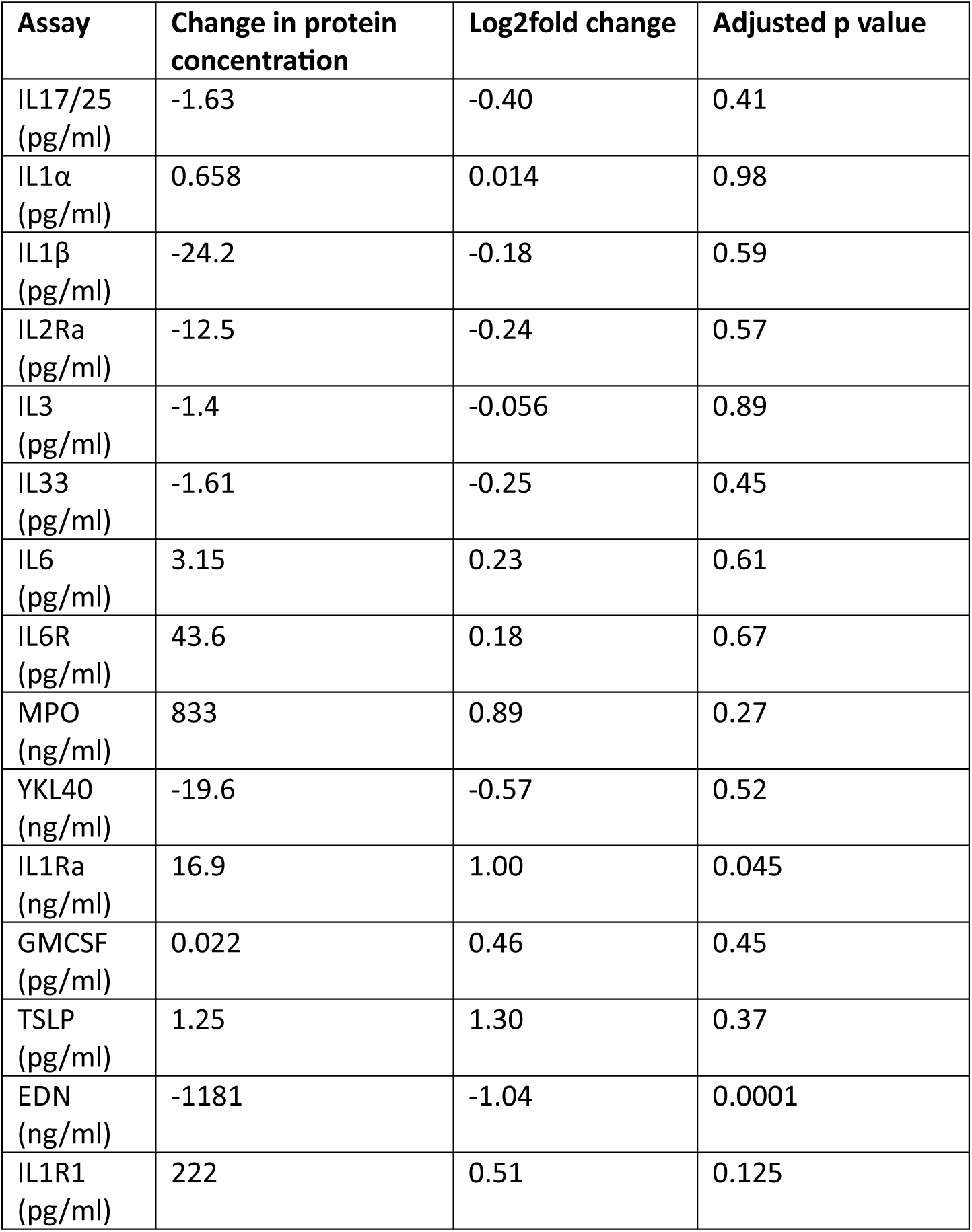
The effect of prednisolone versus placebo on sputum proteins (ELISA)

| **Assay** | **Change in protein concentration** | **Log2fold change** | **Adjusted p value** |
| --- | --- | --- | --- |
| IL17/25 (pg/ml) | -1.63 | -0.40 | 0.41 |
| IL1α (pg/ml) | 0.658 | 0.014 | 0.98 |
| IL1β (pg/ml) | -24.2 | -0.18 | 0.59 |
| IL2Ra (pg/ml) | -12.5 | -0.24 | 0.57 |
| IL3 (pg/ml) | -1.4 | -0.056 | 0.89 |
| IL33 (pg/ml) | -1.61 | -0.25 | 0.45 |
| IL6 (pg/ml) | 3.15 | 0.23 | 0.61 |
| IL6R (pg/ml) | 43.6 | 0.18 | 0.67 |
| MPO (ng/ml) | 833 | 0.89 | 0.27 |
| YKL40 (ng/ml) | -19.6 | -0.57 | 0.52 |
| IL1Ra (ng/ml) | 16.9 | 1.00 | 0.045 |
| GMCSF (pg/ml) | 0.022 | 0.46 | 0.45 |
| TSLP (pg/ml) | 1.25 | 1.30 | 0.37 |
| EDN (ng/ml) | -1181 | -1.04 | 0.0001 |
| IL1R1 (pg/ml) | 222 | 0.51 | 0.125 |

**Supplementary table 8.**
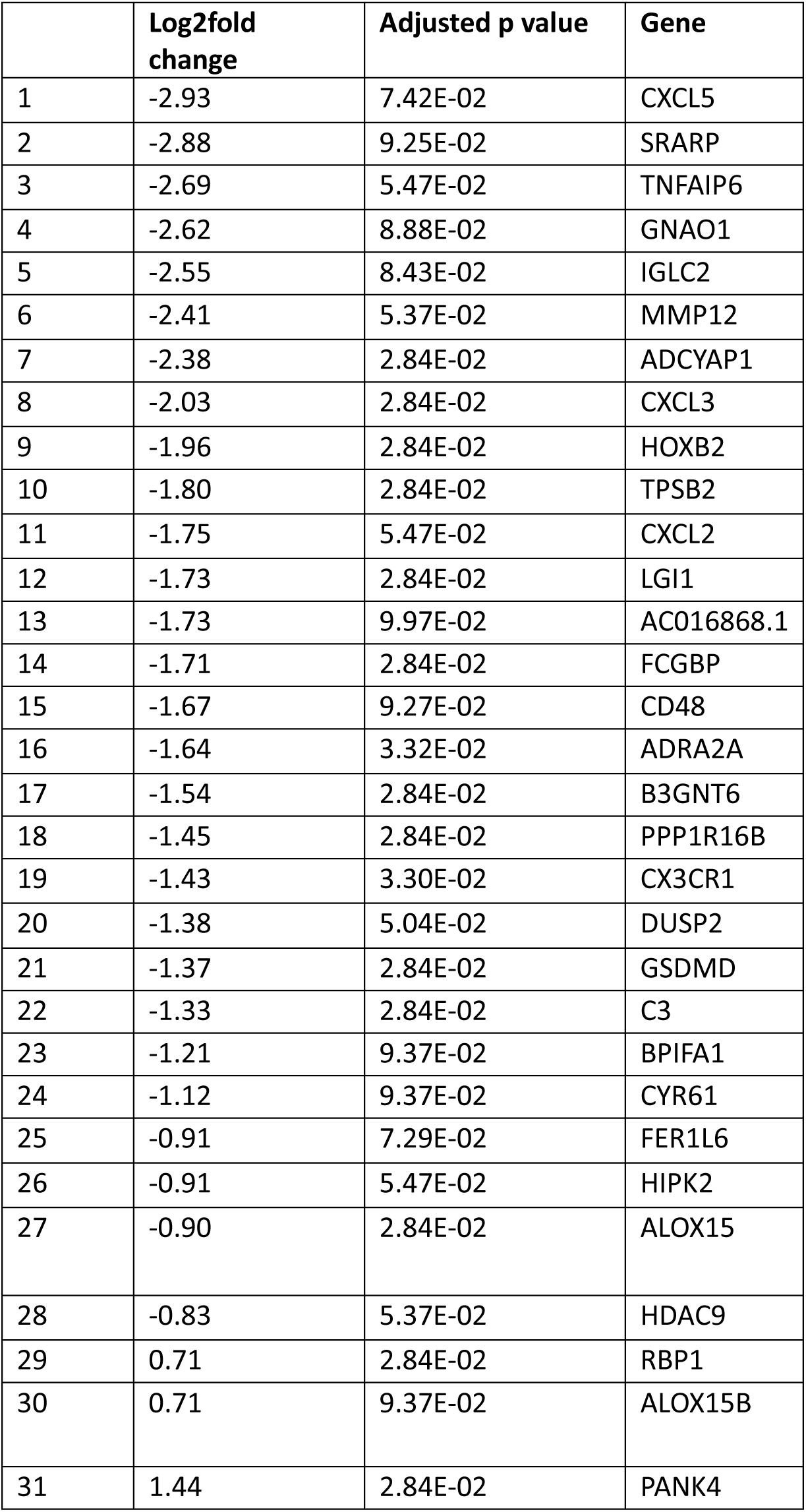
The effect of prednisolone versus placebo on nasal tissue gene transcription.

|  | **Log2fold change** | **Adjusted p value** | **Gene** |
| --- | --- | --- | --- |
| 1 | -2.93 | 7.42E-02 | CXCL5 |
| 2 | -2.88 | 9.25E-02 | SRARP |
| 3 | -2.69 | 5.47E-02 | TNFAIP6 |
| 4 | -2.62 | 8.88E-02 | GNAO1 |
| 5 | -2.55 | 8.43E-02 | IGLC2 |
| 6 | -2.41 | 5.37E-02 | MMP12 |
| 7 | -2.38 | 2.84E-02 | ADCYAP1 |
| 8 | -2.03 | 2.84E-02 | CXCL3 |
| 9 | -1.96 | 2.84E-02 | HOXB2 |
| 10 | -1.80 | 2.84E-02 | TPSB2 |
| 11 | -1.75 | 5.47E-02 | CXCL2 |
| 12 | -1.73 | 2.84E-02 | LGI1 |
| 13 | -1.73 | 9.97E-02 | AC016868.1 |
| 14 | -1.71 | 2.84E-02 | FCGBP |
| 15 | -1.67 | 9.27E-02 | CD48 |
| 16 | -1.64 | 3.32E-02 | ADRA2A |
| 17 | -1.54 | 2.84E-02 | B3GNT6 |
| 18 | -1.45 | 2.84E-02 | PPP1R16B |
| 19 | -1.43 | 3.30E-02 | CX3CR1 |
| 20 | -1.38 | 5.04E-02 | DUSP2 |
| 21 | -1.37 | 2.84E-02 | GSDMD |
| 22 | -1.33 | 2.84E-02 | C3 |
| 23 | -1.21 | 9.37E-02 | BPIFA1 |
| 24 | -1.12 | 9.37E-02 | CYR61 |
| 25 | -0.91 | 7.29E-02 | FER1L6 |
| 26 | -0.91 | 5.47E-02 | HIPK2 |
| 27 | -0.90 | 2.84E-02 | ALOX15 |
| 28 | -0.83 | 5.37E-02 | HDAC9 |
| 29 | 0.71 | 2.84E-02 | RBP1 |
| 30 | 0.71 | 9.37E-02 | ALOX15B |
| 31 | 1.44 | 2.84E-02 | PANK4 |

**Supplementary table 9.** Precursor miRNA significantly differentially expressed by prednisolone versus placebo.

|  | **Pre-cursor miRNA** | **Difference in** | **Adjusted p value** |
| --- | --- | --- | --- |

|  |  | **normalised**<br>**gene count** |  |
| --- | --- | --- | --- |
| 1 | hsa-mir-6511a-3 | -58.8 | 0.00252 |
| 2 | hsa-mir-6511a-2 | -58.8 | 0.00252 |
| 3 | hsa-mir-6511a-1 | -58.8 | 0.00252 |
| 4 | hsa-mir-6511a-4 | -58.8 | 0.00252 |
| 5 | hsa-mir-516a-1 | 111 | 0.00592 |
| 6 | hsa-mir-516a-2 | 111 | 0.00592 |
| 7 | hsa-mir-5701-2 | -77.5 | 0.0114 |
| 8 | hsa-mir-5701-3 | -77.5 | 0.0114 |
| 9 | hsa-mir-5701-1 | -77.5 | 0.0114 |
| 10 | hsa-mir-518a-2 | -84.4 | 0.0405 |
| 11 | hsa-mir-518a-1 | -84.4 | 0.0405 |
| 12 | hsa-mir-3670-2 | 24.0 | 0.0448 |
| 13 | hsa-mir-3670-4 | 24.0 | 0.0448 |
| 14 | hsa-mir-3670-3 | 24.0 | 0.0448 |
| 15 | hsa-mir-3670-1 | 24.0 | 0.0448 |
| 16 | hsa-mir-4520-1 | 44.9 | 0.0687 |
| 17 | hsa-mir-4520-2 | 39.9 | 0.0687 |
| 18 | hsa-mir-4650-2 | -30.9 | 0.0930 |
| 19 | hsa-mir-4650-1 | -30.9 | 0.0930 |

**Supplementary table 10.**
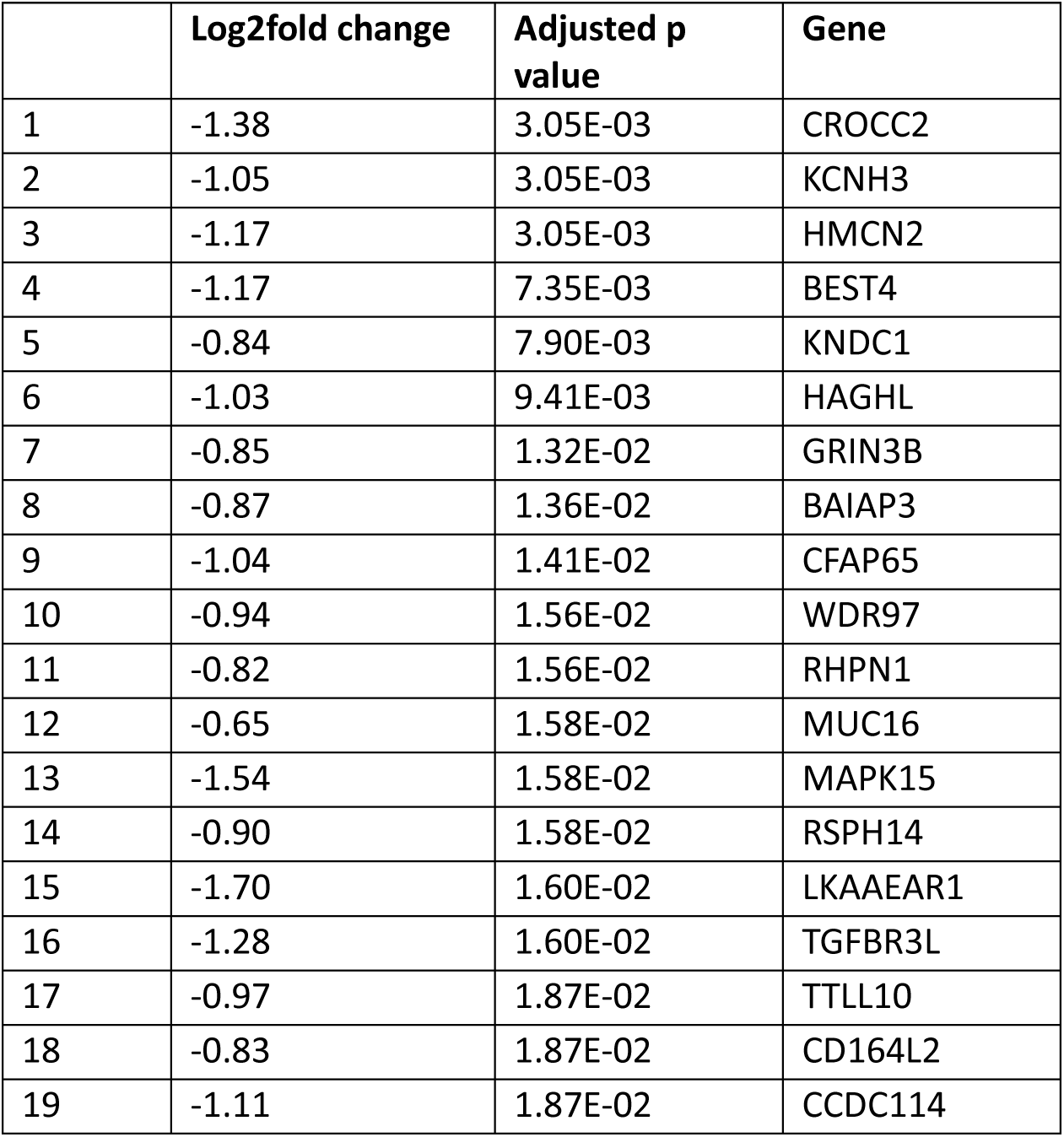
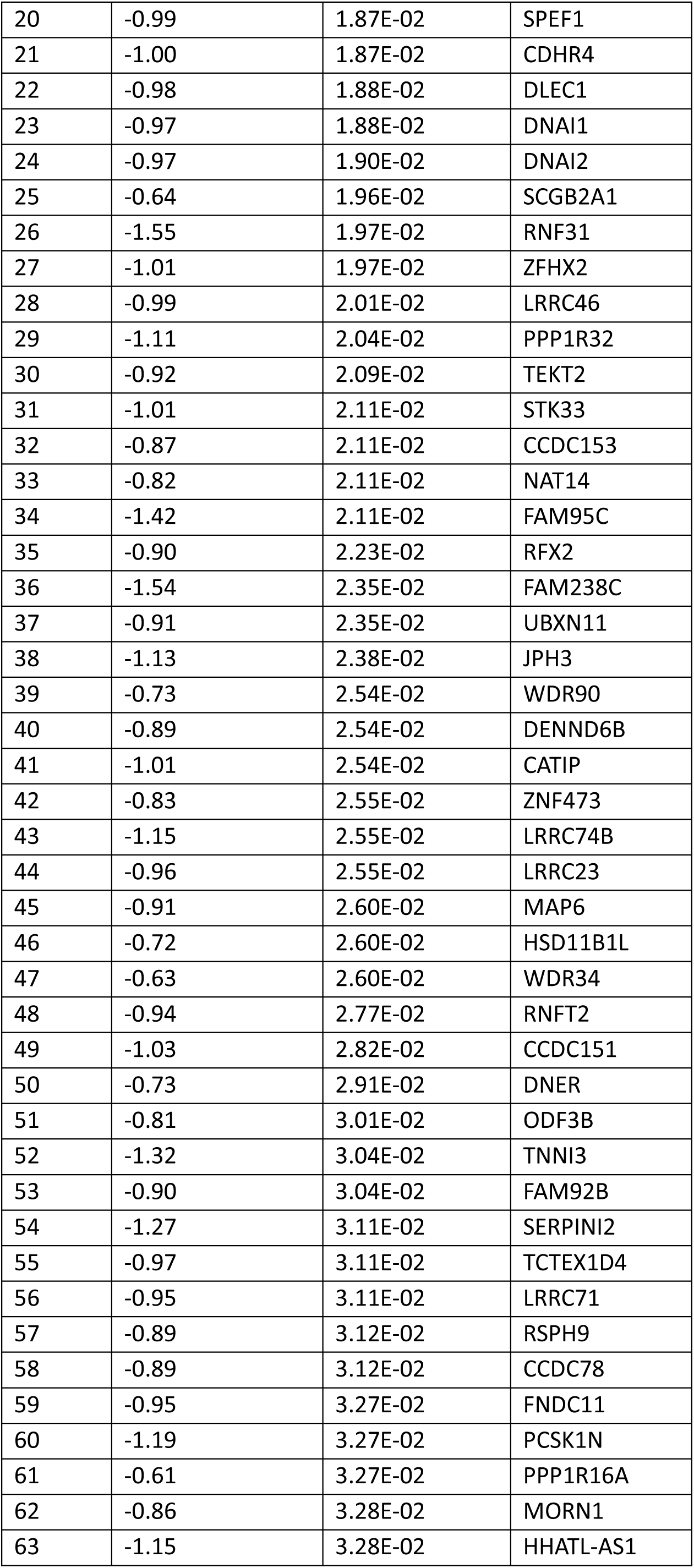
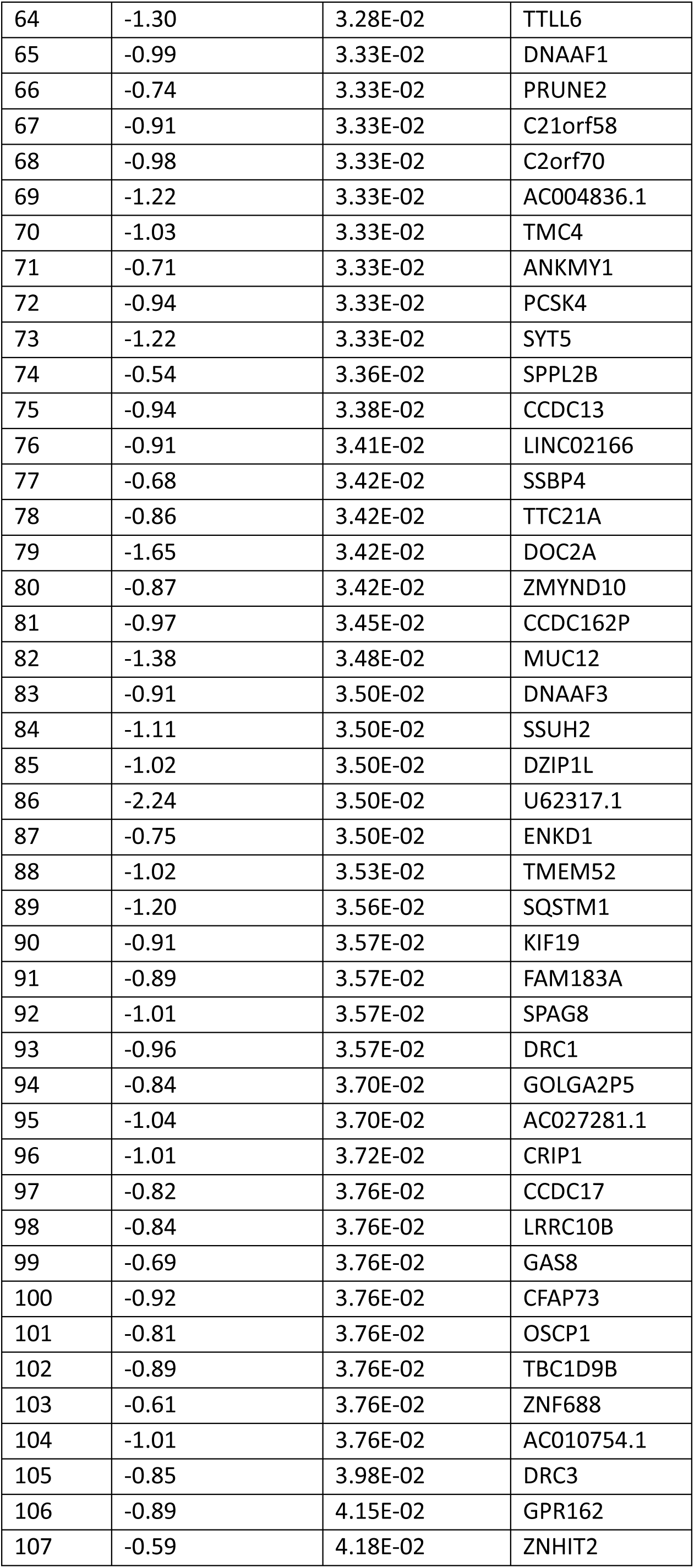
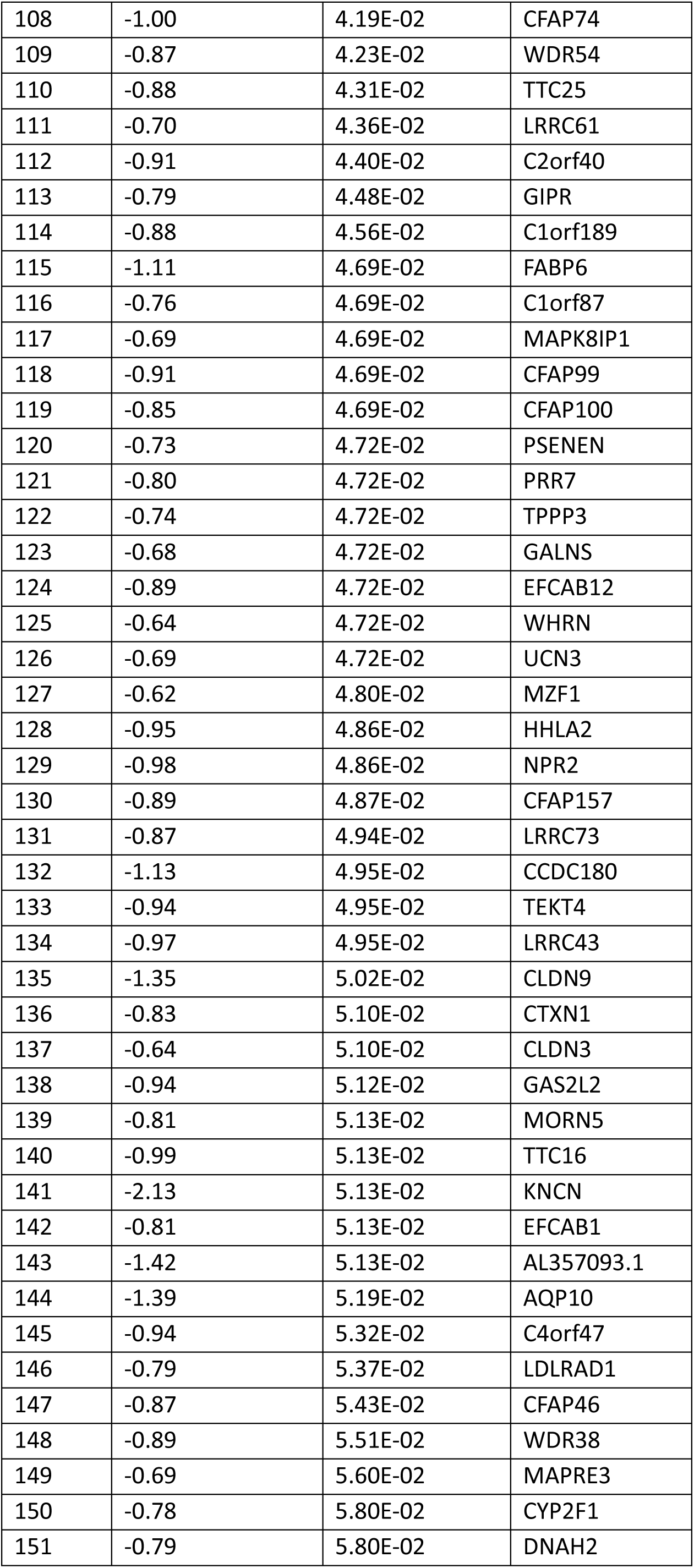
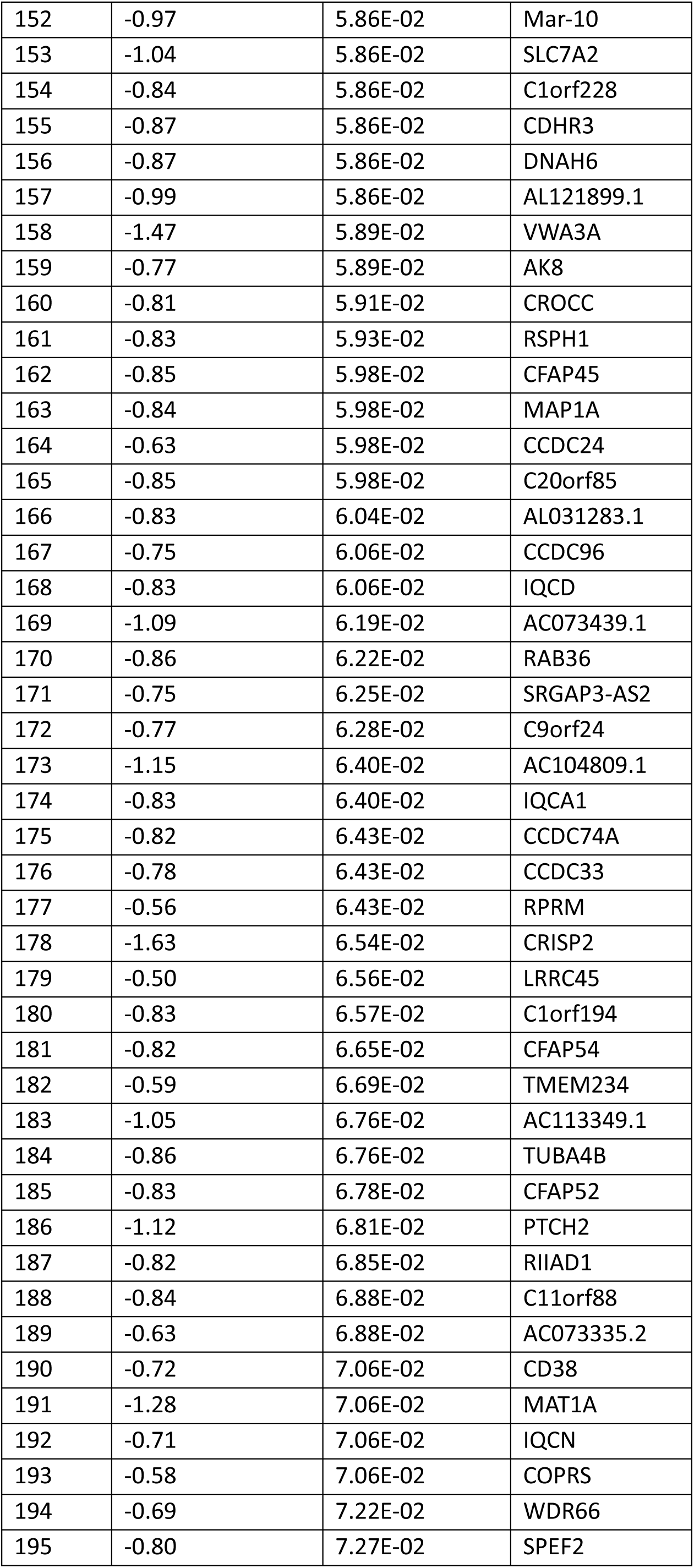
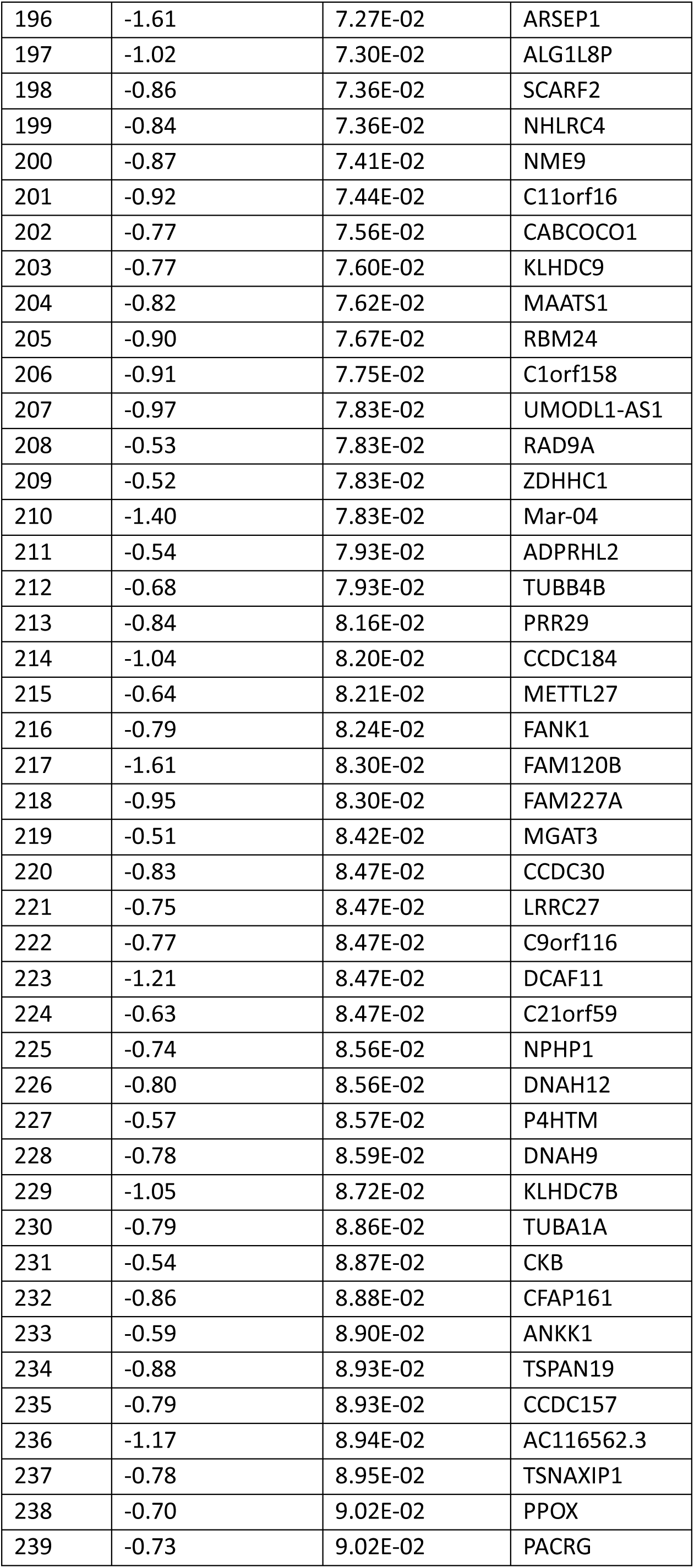
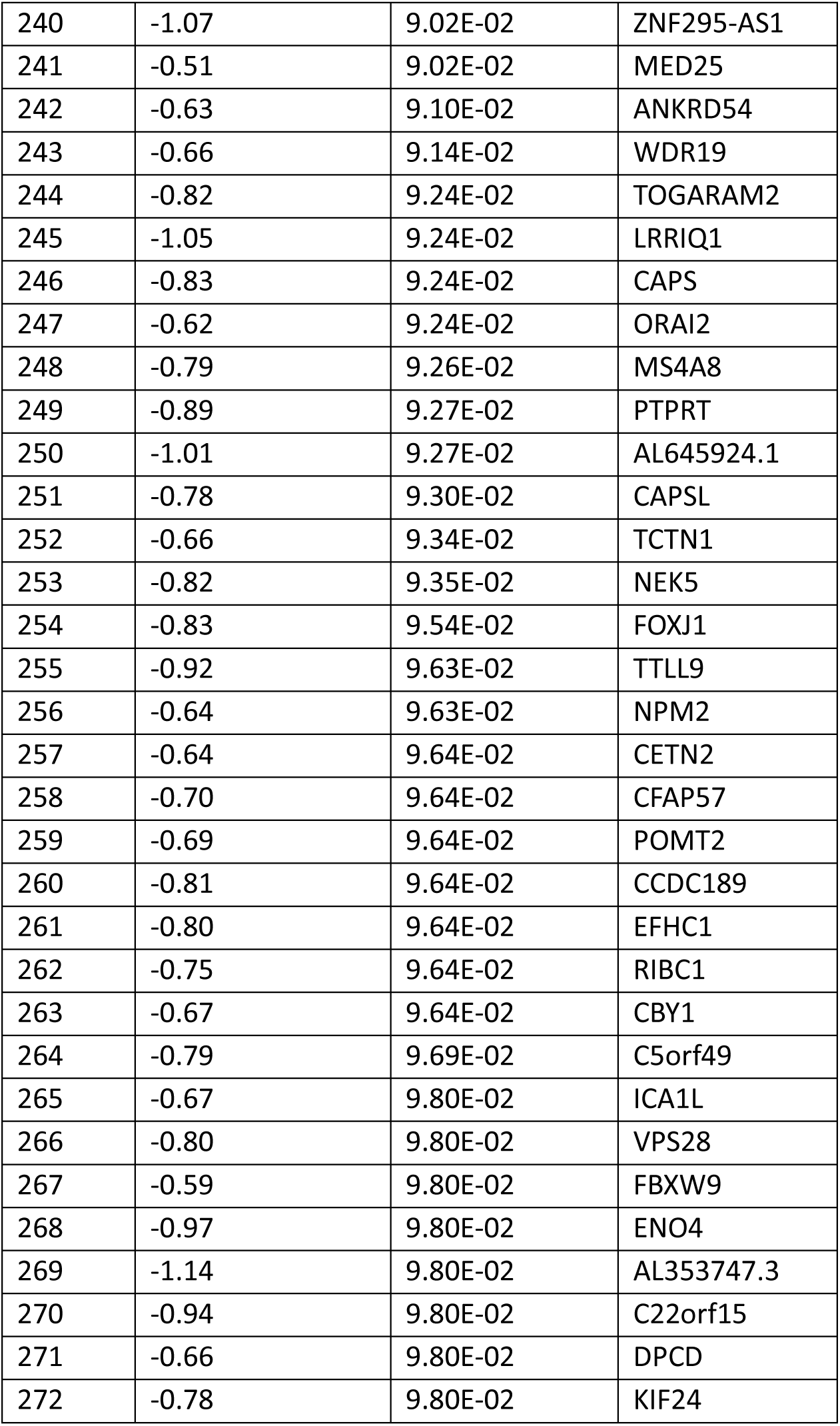
Nasal tissue genes significantly downregulated by mepolizumab.

**Supplementary table 11.**
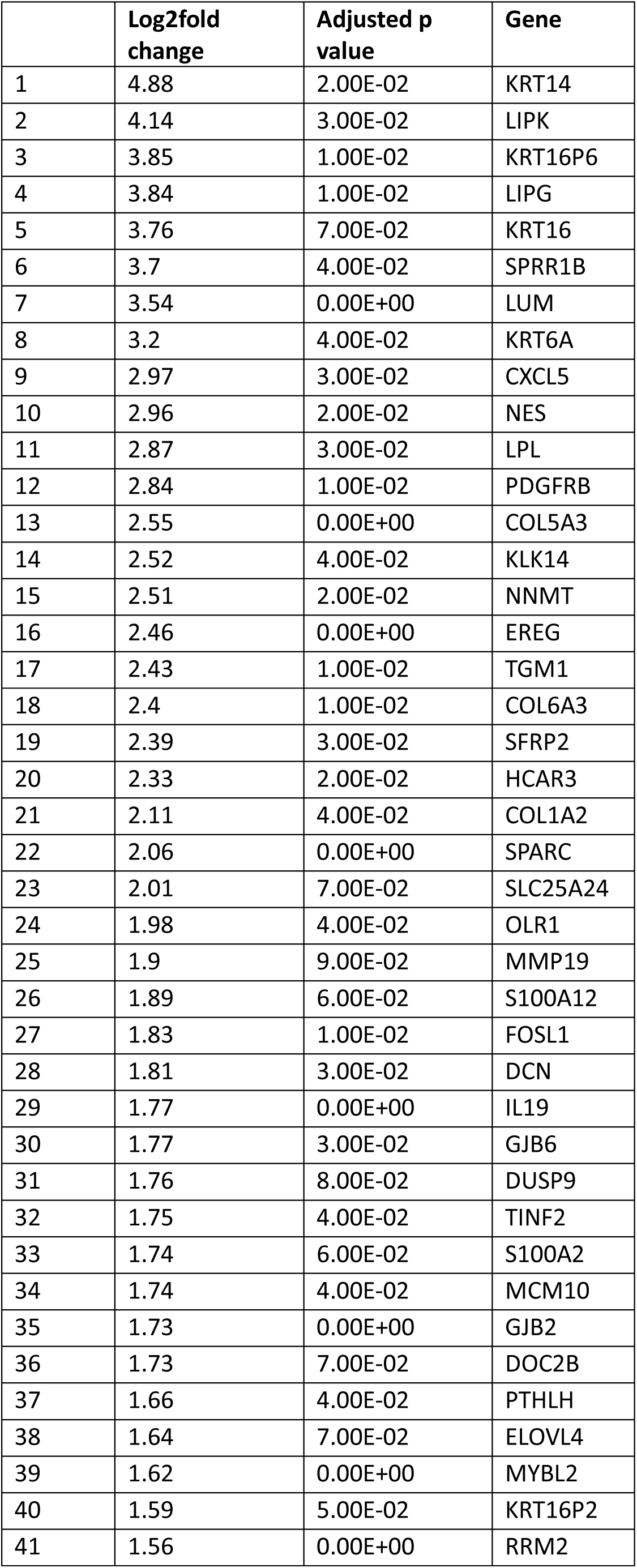
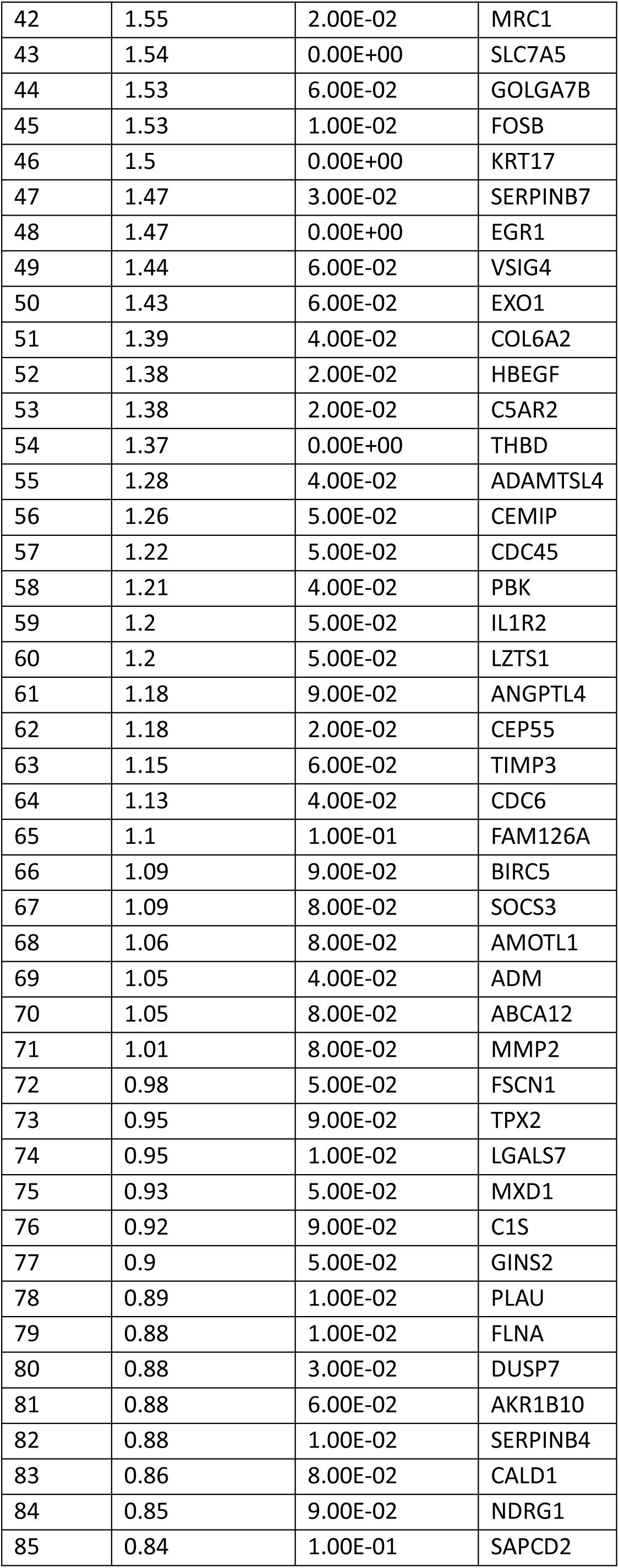
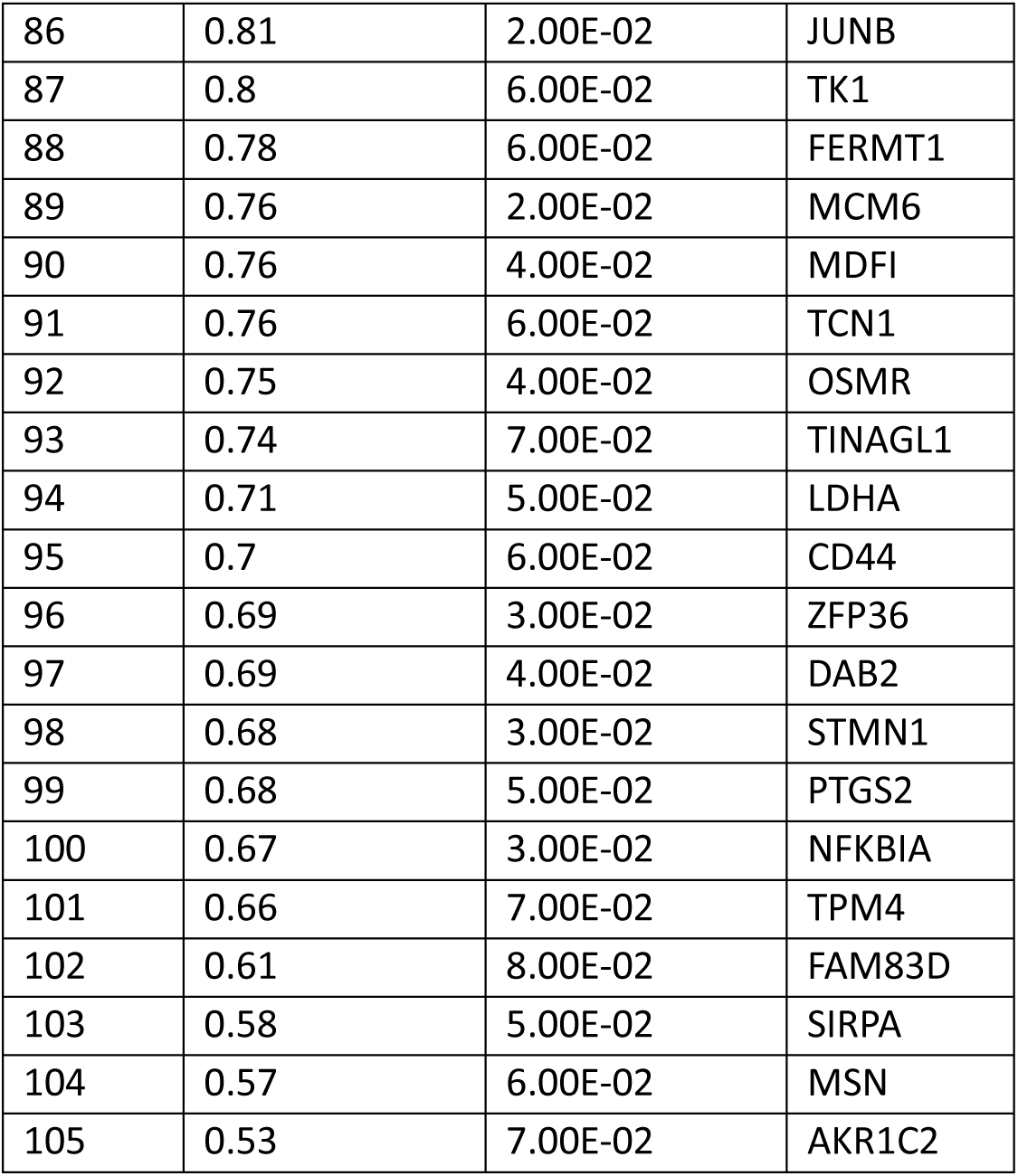
Nasal tissue genes significantly upregulated by mepolizumab.

## Appendix

Summaries of RASP-UK standard operating procedures for sputum induction, nasal scrape sampling, and sputum processing in the MAPLE trial are as follows:

### Sputum induction

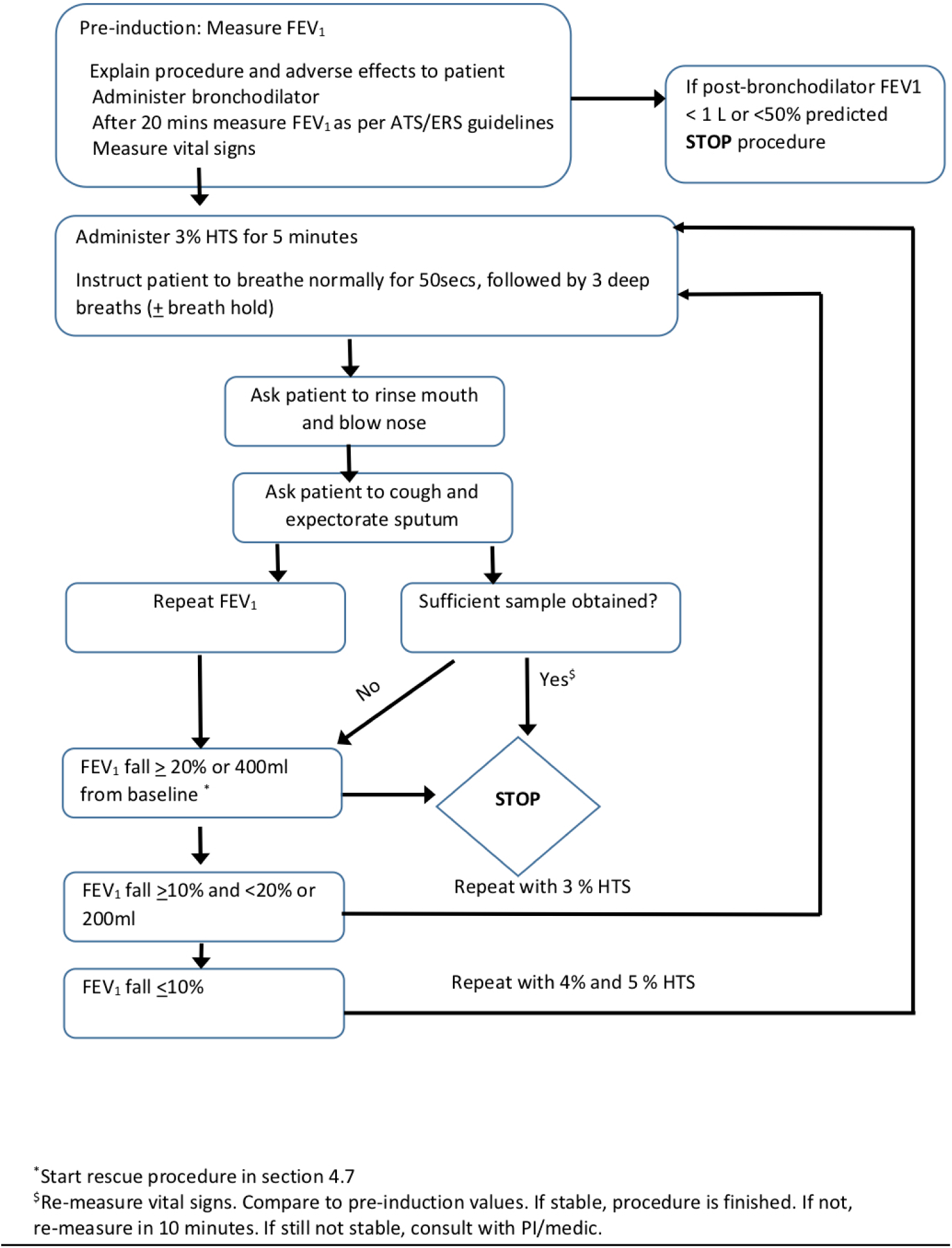

### Nasal scrape sampling

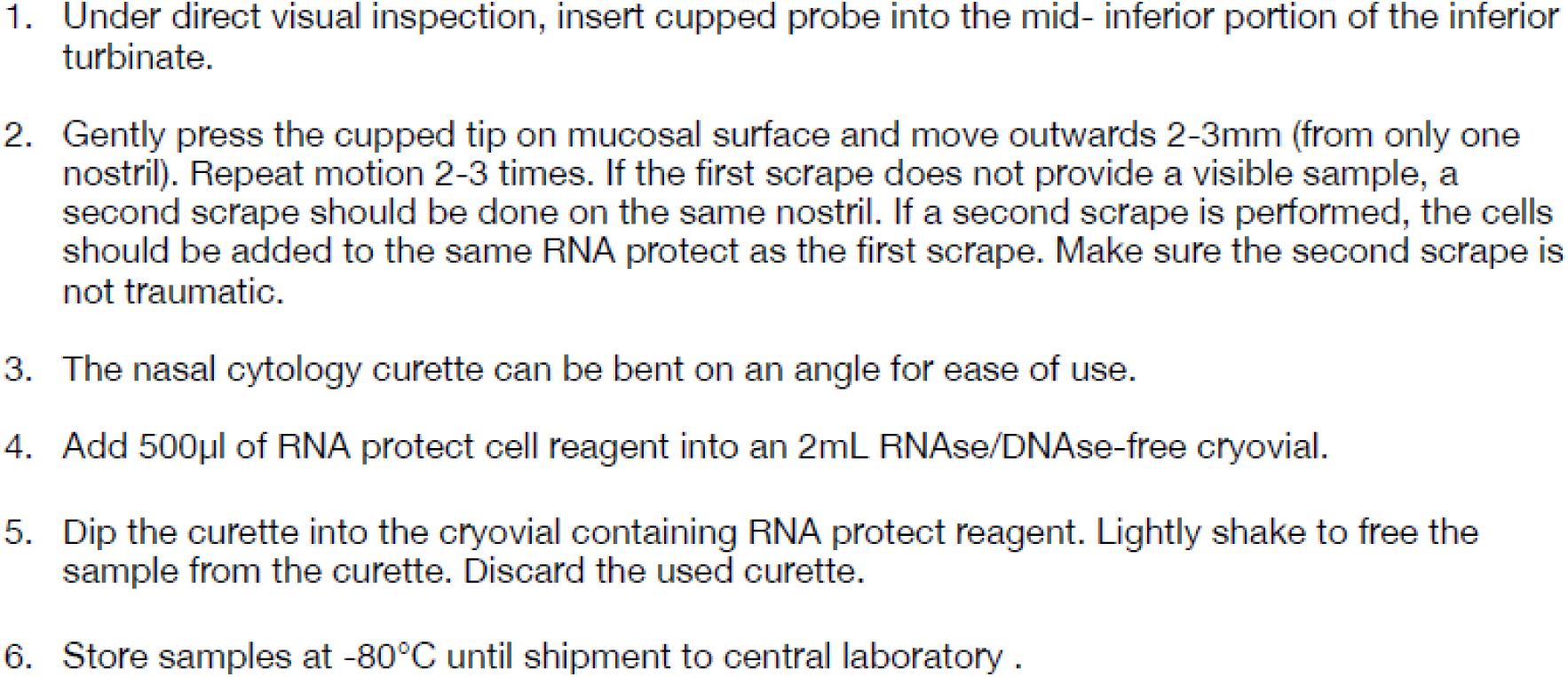

### Sputum processing

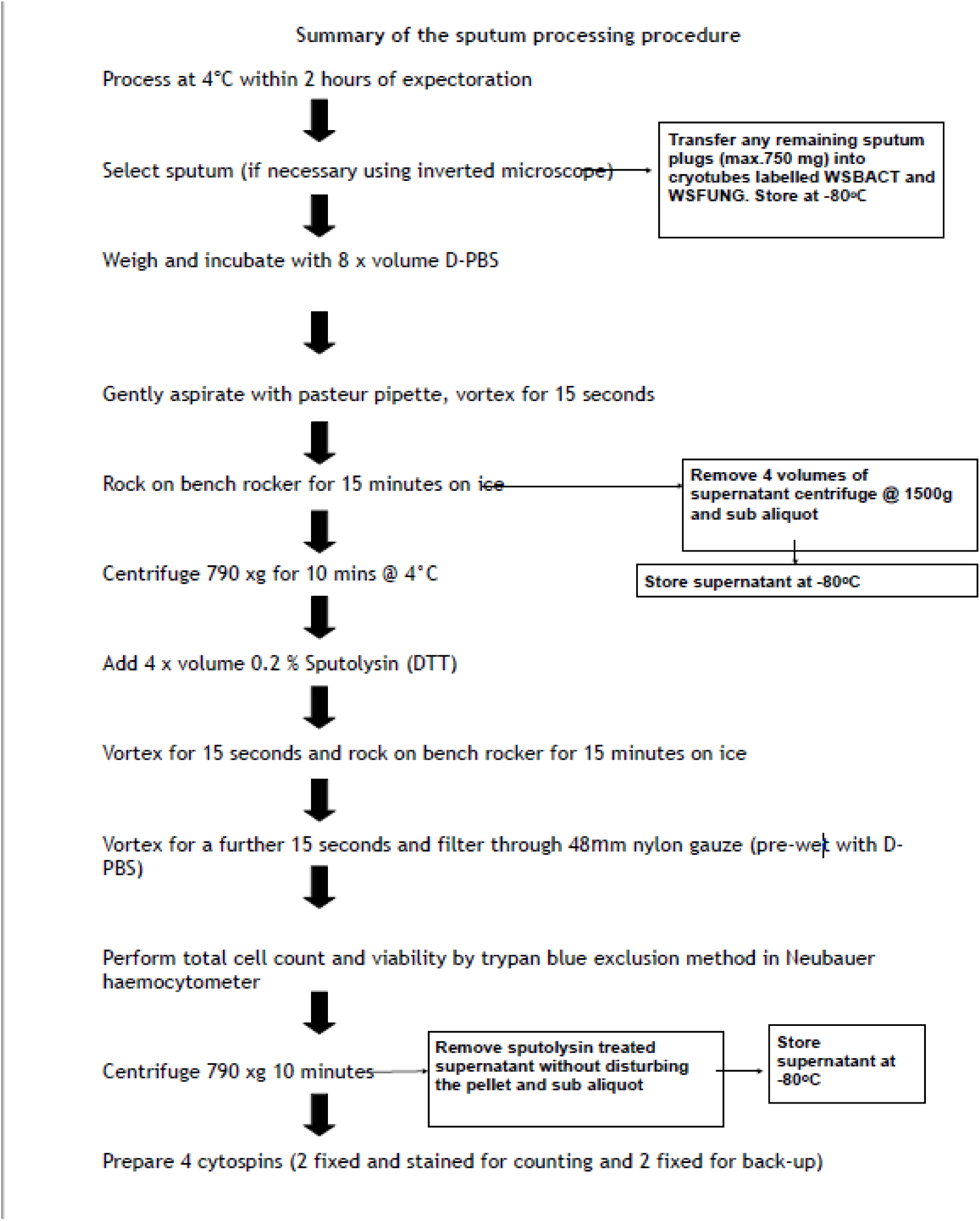

